# Second wave of COVID-19 in India could be predicted with genomic surveillance of SARS-CoV-2 variants coupled with epidemiological data: A tool for future

**DOI:** 10.1101/2021.06.09.21258612

**Authors:** Ashutosh Kumar, Adil Asghar, Prakhar Dwivedi, Gopichand Kumar, Ravi K. Narayan, Rakesh K. Jha, Rakesh Parashar, Chetan Sahni, Sada N. Pandey

## Abstract

India recently witnessed a devastating second wave of COVID-19, which peaked by the end of the first week of May 2021. We aimed to understand formation and spread of the second wave in the country. We analyzed time series distribution of the genomic sequence data for SARS-CoV-2 and correlated that with the epidemiological data for new cases and deaths, for the corresponding period of the second wave. Further we analyzed the phylodynamics of the circulating SARS-CoV-2 variants in the Indian population in the period of study. Our analysis shows that the first indications of arrival of the second wave were observable by the end of January 2021, and by the end of March, 2021 it was clearly indicated. B.1.617 lineage variants drove the wave, particularly B.1.617.2 (a.k.a. delta variant). Based on the observations of this study, we propose that genomic surveillance of the SARS-CoV-2 variants augmented with epidemiological data can be a promising tool for forecasting imminent COVID-19 waves.

## Introduction

India witnessed a devastating second wave of COVID-19 that started towards the end of February 2021. An unwarned arrival of the second COVID-19 wave and the exponential surge in the infections crumpled the country’s epidemic response system and health infrastructure. This resulted in massive suffering and loss of lives, which could have been significantly minimized if timely forecasts were available. A lot of policy discourse has, since then, revolved around whether an imminent COVID-19 wave can be successfully forecasted. The second wave in India was arguably triggered by an emerging lineage of SARS-CoV-2 variants B.1.617, particularly its sub-lineage B.1.617.2 (a.k.a. delta variant) (1). B.1.617 lineage was recently recognized as a global variant of concern (VOC) by World Health Organization. B.1.617 variant was first reported in India in October 2020 (9), and the strain evolved to three more sub-lineages B.1.617.1-3. As per the most recent update from WHO, currently B.1.617.1 (a.k.a. kappa variant) is a variant of interest (VOI) and B.1.617.2 gained status of VOC replacing B.1.617 (2). B.1.617 contained mutations in the key spike protein regions involved in interactions with the host and induction of neutralizing antibodies (S: L452R, E484Q, D614G, del681, del1072) (3). The sub-lineages contained lineage defining spike mutations (L452R, D614G) as well as developed new mutations: B.1.617.1 (S: T95I, G142D, E154K, L452R, E484Q, D614G, P681R, Q1071H), B.1.617.2 (S: T19R, G142D, 156del, 157del, R158G, L452R, T478K, D614G, P681R, D950N), and B.1.617.3 (S: T19R, L452R, E484Q, D614G, P681R) (4). The recent studies suggest that B.1.617 lineage variants are more transmissible (5-10) as well as more lethal (8) than B.1.1.7 (a.k.a. alpha variant), which had been a dominant strain in Indian population before the arrival of second wave (1). The studies have shown a significant reduction in the neutralization against B.1.617 lineage variants by antibodies received from natural infections and many currently used COVID-19 vaccines and multiple monoclonal antibodies (5-8). Higher transmissibility and immunoescape is being reported for B.1.617.2, which is currently the fastest growing SARS-CoV-2 strain in the Indian population (1, 9-10). Until December 2020, B.1.1.7 was a dominant variant in the Indian population, although the first cases of B.1.617 lineage had started appearing by then. By April 2021 India entered a full blown second COVID-19 wave (1, 2). In this study, we analyzed the viral genomic surveillance and epidemiological data to ascertain the causative SARS-CoV-2 strain and to delineate its spread in the population in a time course leading to second COVID-19 wave in India.

## Methods

### (Study design, participants, data sources, and statistical analyses)

A time series (weekly and monthly) distribution of the genomic sequence data for SARS-CoV-2 from India along with the epidemiological data for new cases and deaths from COVID-19 were analyzed for the period from 1^st^December 2020 to 26^th^ July 2021 (a total of 34 weeks). Further, the phylodynamic analysis of the individual variants circulating in the population in the period of study was performed.

### Weekly and monthly distribution of SARS-CoV-2 variants and new cases and deaths due to COVID-19

The genomic sequence data for SARS-CoV-2 and official epidemiological data for COVID-19 for the period from 1^st^ December 2020 to 26^th^ July 2021 from India were downloaded from EpiCoV^™^ database of Global Initiative on Sharing All Influenza Data (GISAID) and Worldometer: https://www.worldometers.info/coronavirus/coronavirus/country/india/) respectively. A total of 40359 SARS-CoV-2 genomic sequences were analyzed. The number of sequences for each SARS-CoV-2 variants was retrieved using automatic search function feeding information for the lineage/sub-lineage and collection dates in EpiCoV^™^ database of GISAID. Total number of sequence per week and month for the studied time period was noted for each variant and their relative proportions were calculated (in percent). Data was tabulated, and weekly and monthly distribution of each variant was charted against the COVID-19 epidemiological data (new case and deaths) and statistically analyzed. The graphs were plotted to visualize the trends. Further, the genomic sequence of SARS-CoV-2 variants were analyzed for the individual states and union territories, to check if there has been any deviation from the collective data trends.

### Phylodynamics of SARS-CoV-2 variants

A phylodynamic analysis of the variants circulating in the Indian population in the period of study was performed from GISAID sequences using bioinformatics tool available at EpiCoV^™^.

### Statistical analysis

All statistical analyses were performed using XLSTAT. Descriptive statistics was calculated for all of the variables. Normal distribution and homogeneity of the data were determined using Andersons Darling and Levene’s tests, respectively. A correlation matrix was generated and further a linear regression analysis was conducted between the comparing variables (presented as R values= 0 to 1). For all the comparisons statistical significance threshold was set as p ≤ 0.05.

### Ethics approval

An approval from the institute ethics committee was precluded as the data used in this study were retrieved from publically available databases.

### Funding

No substantial funding was received for this study.

## Results and Discussion

A retrospective analysis of the epidemiological data reflected that the second COVID-19 wave started rising by the end of February, 2021 and peaked by the end of the first week of May 2021. Based on the distinct epidemiological trends observable in the graph (Fig. S1), we divided the period of study (1^st^ December 2020 to 26^th^ July 2021, a total of 34 weeks) into pre peak (1-23 week) and post peak (24-34 week) period (Fig. S1). The weekly average of new cases and deaths showed strong correlation in the complete study period (R=0.98, p<.001) signifying high statistical validity of the data for the further comparisons. Further, we analyzed the distribution of SARS-CoV-2 variants circulating in the Indian population in correlation of the new cases and deaths in the period before the peak and thereafter. For the purpose of description, based on the epidemiological trends, the pre peak period was further divided into three time series intervals: ‘very early’ (1-8 weeks), ‘early’ (9-16 weeks), and ‘near peak’ (17-23 weeks). In the ‘very early’ period new cases and deaths showed a down ward trend, which maintained a plateau in the ‘early’ period (except towards the end when cases and deaths started rising indicating start of the second wave). In the ‘near peak’ period steep rise in the new cases and deaths were observable (Fig. 1).

**Figure 1.**
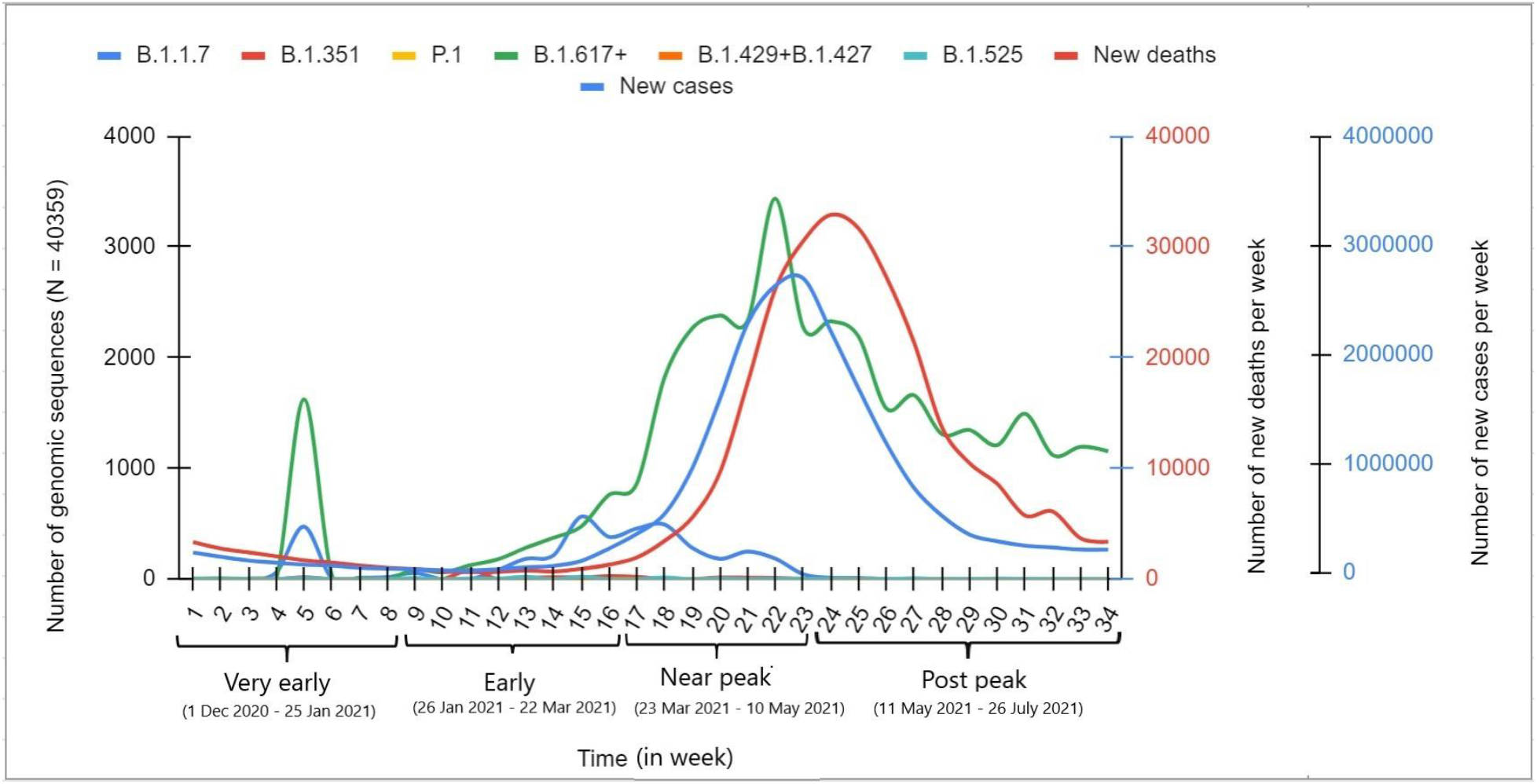
Weekly distribution of SARS-CoV-2 variants in genomic sequence data from India and their correlation with daily new COVID-19 cases and deaths for the period of 1st December 2020 to 26th July 2021. [The data were analyzed for the period before the peak of the second wave (23^rd^ week) and thereafter. For the purpose of description, based on the epidemiological trends, the pre peak period was further divided into three time series intervals: ‘very early’ (1-8 weeks), ‘early’ (9-16 weeks), and ‘near peak’ (17-23 weeks). (Data source: SARS-CoV-2 genomic sequence—GISAID database: https://www.gisaid.org/; Epidemiological data—Worldometer: https://www.worldometers.info/coronavirus/coronavirus/country/india.)]

The rise and fall of the circulating SARS-CoV-2 variants were studied against the observed epidemiological data trends in the respective time series intervals. Observing the composite data trends of epidemiological and SARS-CoV-2 genomic data gives a glimpse of the formation of the second COVID-19 wave in with clear indications which SARS-CoV-2 strains may have driven it (Figs. 1-2). By December 2020, eight SARS-CoV-2 pango lineages and their multiple sub-lineages were circulating in Indian population including four VOCs (B.1.1.7, B.1.351, P1, and B.1.617.2) and three variants of interest (VOIs) (B.1.617.1, B.1.127/B.1.429 and B.1.525). However, B.1.1.7 was the most dominant variant in that period. B.1.617 lineage variants collectively (B.1.617+) showed an upward trend since the emergence and surpassed other VOCs including B.1.1.7 by end of January 2021 (8-9^th^ week), and then kept rising up thereafter. In contrast, B.1.1.7 showed a downward trend by the end of March 2021 (17-18^th^ week). This indicated B.1.617 lineage variants were the dominant variant hereafter. Confirming to this trend, by the end of April 2021 B.1.617 lineage variants were detected in 78.5 % of SARS-CoV-2 sequences uploaded on GISAID, and that further reached to about 83% in the week of peak.

The rise of B.1.617 lineage variants showed an evolution in the relationship with the epidemiological data, showing weak to strong statistical correlation with the rise of new cases, in ‘very early’ (R= 0.36, p> .05) and ‘early’ (R= 0.26, p> .05) to ‘near peak’ (R= 0.87, p= 0.01) time interval; which indicated intensification of the second wave as the relative proportion of these variants increased (Fig. 1). Of note, this trend was not observed with any other variant, signifying that the B.1.617 lineage was the prime driver of the rising second wave. Further, we wanted to know which particular B.1.617 lineage variants were dominating in the studied time period. Interestingly, an intra-lineage competition was distinctly visible between the sub-lineages of B.1.617 (Fig. 2). The single sample of B.1.617 was reported on 25. 02. 2021 (date of collection) (EPI-ISL_1544002) and thereafter it has not been detected. First case of B.1.617.2 was detected as early as 21^st^ November, 2020 (EPI_ISL_2373501) followed by on 1^st^ December 2020 (EPI_ISL_1372093), and B.1.617.3 on 14^th^ December 2021 (EPI_ISL_2099648). Up to end of February (week 13), B.1.617.1 was detected in higher number of sequences than B.1.617.2, which was followed by B.1.617.3. However by the end of March (week 18) B.1.617.1 took a downturn, but B.1.617.2 continued to rise along with similar rise in the number of new cases. Notably, by the end of April 2021 B.1.617.2 was singly detected in about 72% of the SARS-CoV-2 samples from India uploaded in GISAID database that further reached to about 79% in the week of peak (23^rd^ week). Notably, B.1.617.2, but no other B.1.617 lineage variants, matched the rise in the new cases leading to the peak of wave (Fig. 2) (very early: R= 0.35, p> .05, early: R=0.17, p> .05; and near-peak: R=0.92, p=.0028) (Table S1).

**Figure 2.**
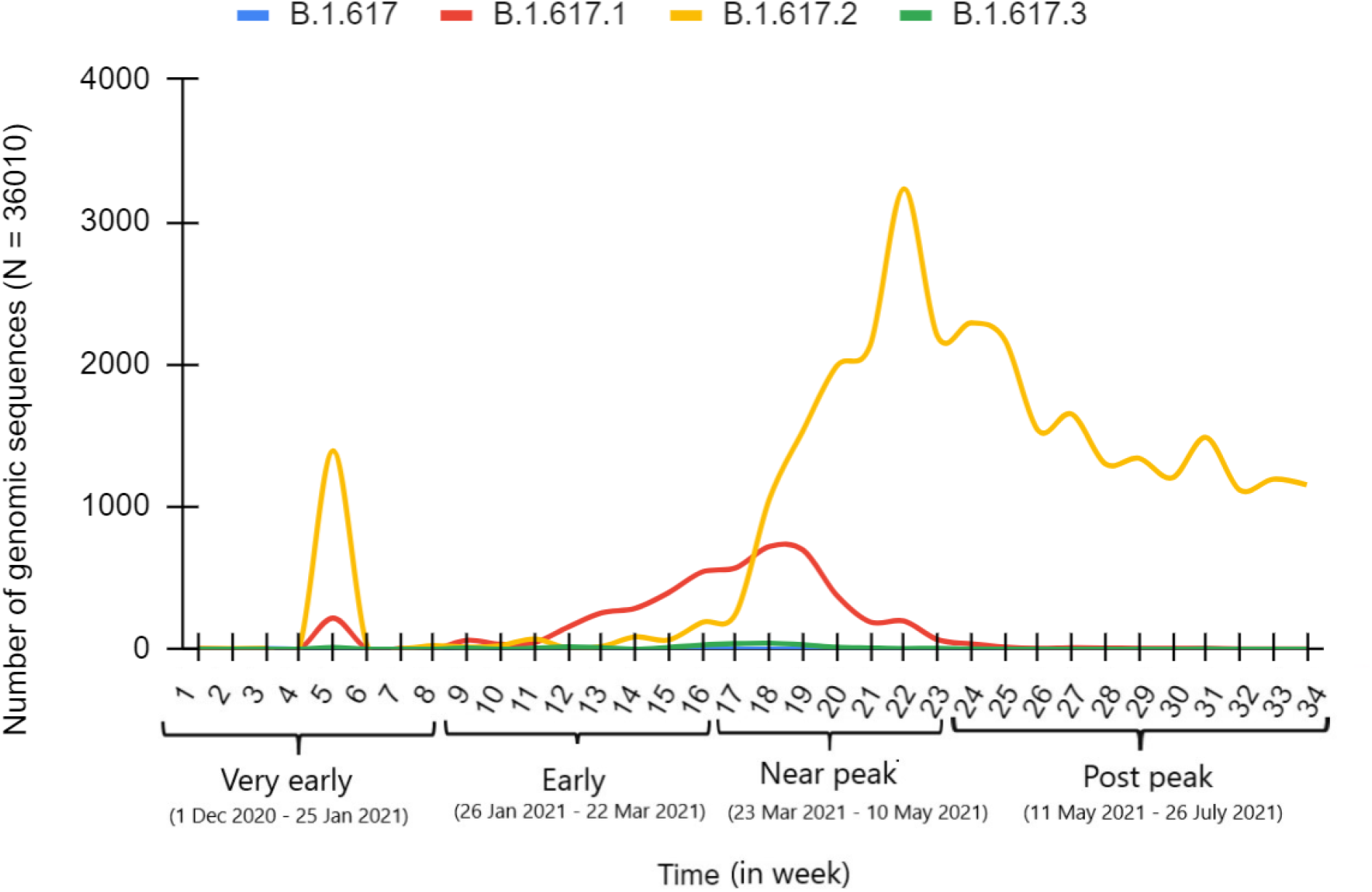
Origin and spread of B.1.617 lineage SARS-CoV-2 variants in Indian population. (Data source: SARS-CoV-2 genomic sequence—GISAID database: https://www.gisaid.org. Data was analyzed for the period of 1^st^ December, 2020 to 26^th^ July 2021.)

The phylodynamic analysis of the circulating variant in the period of study strongly corroborated with the trends presented in the graphs, showing an exclusive increase in the cluster density of in comparison to the other variants in the near-peak period (Fig. 3).

**Figure 3.**
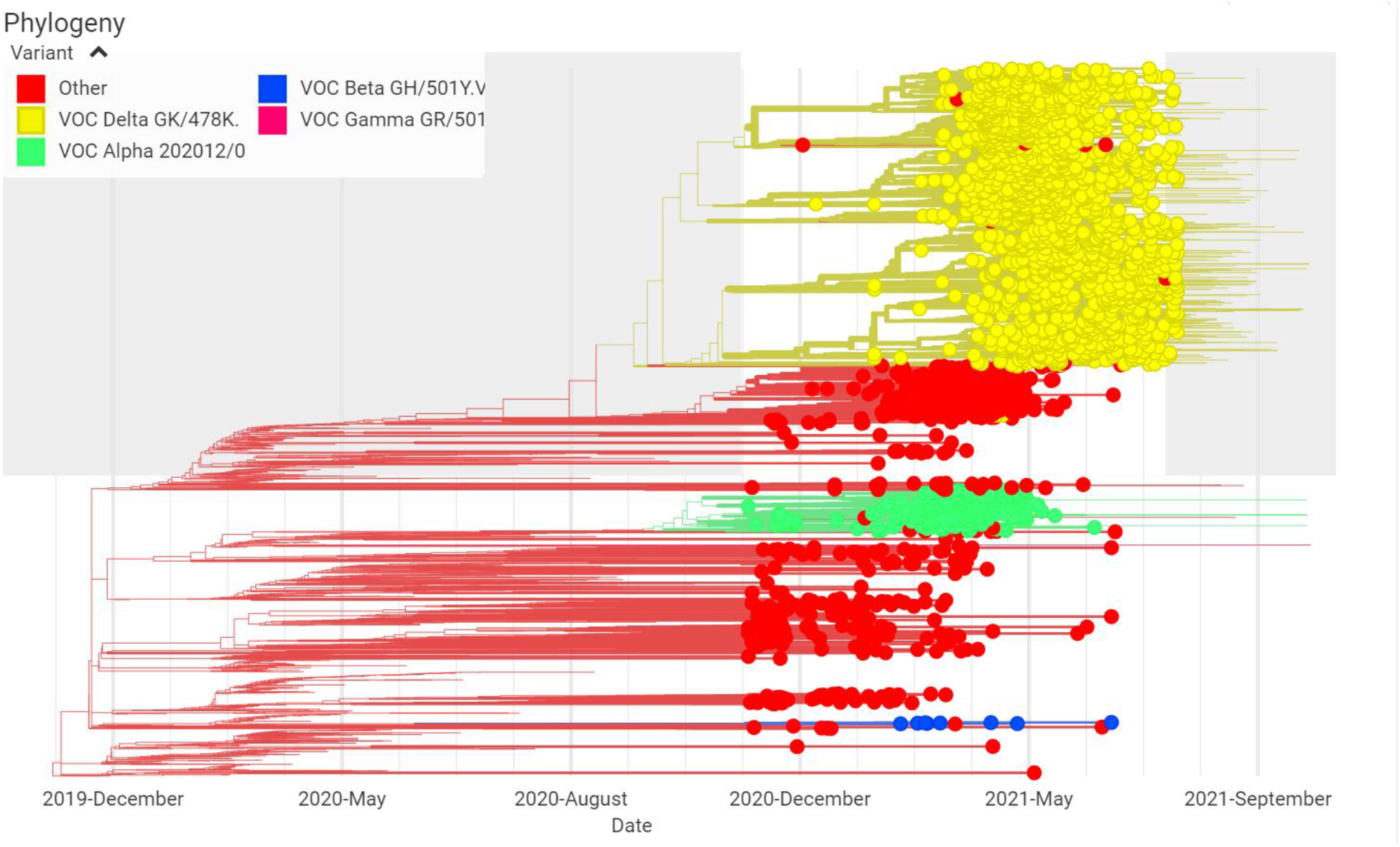
Phylodynamics of SARS-CoV-2 variants in Indian population for the period of 1^st^ December 2020 to 26^th^ July 2021. (Data source: SARS-CoV-2 genomic sequence—GISAID database: https://www.gisaid.org.)

To know whether the rise in B.1.617.2 variant was localized to certain geographical regions which may have influenced the collective data trends, we compared the monthly distribution of genomic sequences of the SARS-CoV-2 variants for the states and union territories of the country individually. A similar increase in the detection of B.1.617.2 variant was observable in most of the states and union territories (Fig. S2), except Kerala, where a different patterns was visible (Fig. S2-O). In Kerala rise of B.1.617.2 was slower in comparison to rest of the country (55.5% vs. 72% of total cases by the end of April 2021), which was further confirmed in the state-wise sero-survey data from the period of second wave (44.4% vs. 67.7% of national average) (11). Notably, an intensive rise in the B.1.617.2 cases were observed in Kerala in a later period.

Thus, the analyses collectively distinctly delineate that the formation of the second COVID-19 wave in India was closely associated with the rise of B.1.617 lineage variants, particularly its sub-lineage B.1.617.2. The first indication of an imminent COVID-19 wave was observable by the end of January 2021 when cases by B.1.617.2 surpassed all other variants, and the rise of the wave was clearly observable by the end of March 2021 when cases by B.1.617.2 showed a steep rise matched with the total new cases.

Our findings get corroborated by a recent article published by a group of scientists affiliated with INSACOG—Indian SARS-CoV-2 Genomic Consortia, who observed a similar pattern in rise of B.1.617 lineage, primarily B.1.617.2 variant in Delhi before second wave (1).

The findings of this study signify that the genomic surveillance of the SARS-CoV-2 variants augmented with epidemiological data can be a potential tool for forecasting imminent COVID-19 waves. Nevertheless, the accuracy of the prediction would largely dependent on the population matched viral genomic sequencing and consistency in uploading of the data from all geographical regions, as well as accurate reporting of the epidemiological data, which currently seems a big hindrance restricting timely predictions.

Plausibly, the exclusive rise in the proportion of an emerging SARS-CoV-2 variant matched with the concomitant rise in new cases should inform arrival of a new COVID-19 wave. However, apart from these, considering the other epidemiological factors, such as previous exposure with the related viral strains and immunization status of the population will be necessary to determine the extent of an imminent wave (12). Notably, first COVID-19 wave in India was limited in the extent as were indicated by the sero-survey data, and also very limited population was vaccinated at the beginning of 2021; with emergence of a new variant both of these factors may have created an optimum condition for the rising of a massive second wave. Further, the preventive measures in place, such as lockdowns or restrictions against gatherings, and use of face masks can also influence the prospects and extent of a new wave.

There have been multiple limitations in our study which could have impacted the interpretation of the findings. Firstly, the samples used in our analyses were not representative for populations as for many geographical regions that has been greatly disproportioned. Hence, the genomic sequence data presented in this study doesn’t necessarily reflect the accurate epidemiological scale of spread of the variants in the reported geographical regions, but only shows their relative proportion in the samples for which genomic sequences were uploaded in GISAID database. We assumed that similar proportions between variants exist in the actual population. Secondly, there has been inconsistency in reporting and uploading of the genomic sequences, which constrained examining a daily trend in the spread of the variants. The paucity of the genomic sequences and inconsistency in their uploading on the used databases for some states/union territories made the determination of the variant dominance difficult.

### Data sharing

Primary data used for this study are publicly available on: SARS-CoV-2 genomic sequence— GISAID database: https://www.gisaid.org/; Epidemiological data—Worldometer: https://www.worldometers.info/coronavirus/coronavirus/country/india). The categorized data for the study period can be availed from the corresponding author on reasonable request.

## Data Availability

Data available from corresponding author on reasonable request.

## Acknowledgements

The study has used SARS-CoV-2 genomic sequence and epidemiological data from GISAID (https://www.gisaid.org/) and Worldometer (https://www.worldometers.info/coronavirus/coronavirus/country/india), respectively.

## Author (s) contributions

AK, PD, GK collected samples and analyzed data. AK wrote first draft. AA performed statistical analysis. AK, RKN, RKJ, RP, CS, and SNP reviewed and edited the paper. All authors consented for submitting final draft.

## References

1. Dhar MS, Marwal R, Radhakrishnan VS, et al. (2021) Genomic characterization and epidemiology of an emerging SARS-CoV-2 variant in Delhi, India. Science. eabj9932. doi:10.1126/science.abj9932

2. WHO, Tracking SARS-CoV-2 variants. https://www.who.int/en/activities/tracking-SARS-CoV-2-variants/. Accessed 16 October 2021.

3. Alaa Abdel Latif, Julia L. Mullen, et al. B.1.617 Lineage Report. outbreak.info, available at https://outbreak.info/situation-reports?pango=B.1.617. Accessed 16 October 2021.

4. WHO, Weekly epidemiological update on COVID-19 -11 May 2021. https://www.who.int/publications/m/item/weekly-epidemiological-update-on-covid-19---11-may-2021

5. Tada T, Zhou H, Dcosta BM, et al. (2021) The Spike Proteins of SARS-CoV-2 B. 1.617 and B. 1.618 Variants Identified in India Provide Partial Resistance to Vaccine-elicited and Therapeutic Monoclonal Antibodies. bioRxiv. https://doi.org/10.1101/2021.05.14.444076

6. Hoffmann M, Hofmann-Winkler H, Krüger N, et al. (2021) SARS-CoV-2 variant B. 1.617 is resistant to Bamlanivimab and evades antibodies induced by infection and vaccination. Cell Rep. 36(3):109415. doi:10.1016/j.celrep.2021.109415.

7. Ferreira I, Datir R, Papa G, et al. (2021) SARS-CoV-2 B. 1.617 emergence and sensitivity to vaccine-elicited antibodies. bioRxiv. https://doi.org/10.1101/2021.05.08.443253

8. Yadav, P.D. et al. (2021) SARS CoV-2 variant B. 1.617. 1 is highly pathogenic in hamsters than B. 1 variant. bioRxiv. https://doi.org/10.1101/2021.05.05.442760

9. Planas D, Veyer D, Baidaliuk A, et al. (2021) Reduced sensitivity of infectious SARS-CoV-2 variant B. 1.617. 2 to monoclonal antibodies and sera from convalescent and vaccinated individuals. Nature. Nature. doi: 10.1038/s41586-021-03777-9.

10. Bernal JL, Andrews N, Gower C, et al. (2021) Effectiveness of COVID-19 vaccines against the B. 1.617. 2 variant. medRxiv.https://doi.org/10.1101/2021.05.22.21257658

11. Rajib Disrupt. (2021) After India’s brutal coronavirus wave, two-thirds of population has been exposed to SARS-CoV2. The Conversation. https://theconversation.com/after-indias-brutal-coronavirus-wave-two-thirds-of-population-has-been-exposed-to-sars-cov2-165050. Accessed 16 October 2021.

12. Dyson L, Hill EM, Moore S, et al. (2021) Possible future waves of SARS-CoV-2 infection generated by variants of concern with a range of characteristics. Nat Commun. 12(1):5730. doi:10.1038/s41467-021-25915-7

