## Supplementary material for "Second wave of COVID-19 in India could be predicted with genomic surveillance of SARS-CoV-2 variants coupled with epidemiological data: A tool for future": Fig. S1, and Fig. S2

**Kumar et al., 2021**

**Supplementary files:**

**Contents: Figure S1 and S2**

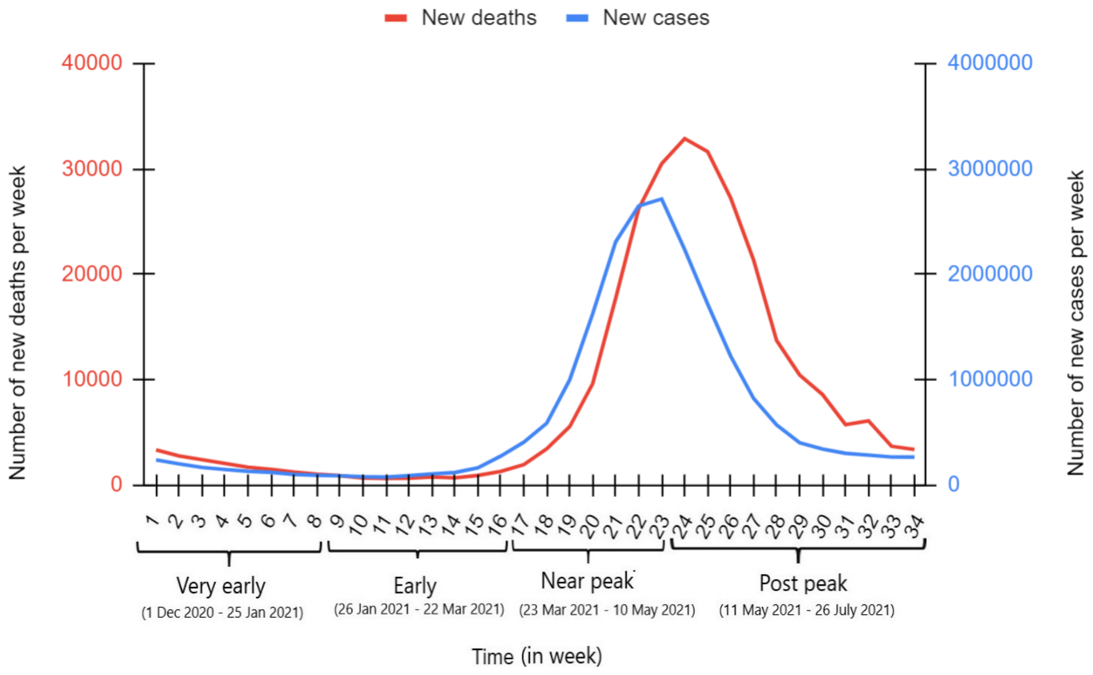


**Figure S1 Weekly new COVID-19 cases and deaths in Indian population for the period of 1st December 2020 to 26th July 2021.** The data were analyzed for the period before the peak of the second wave (23rd week) and thereafter. For the purpose of description, based on the epidemiological trends, the pre peak period was further divided into three time series intervals: ‘very early’ (1-8 weeks), ‘early’ (9-16 weeks), and ‘near peak’ (17-23 weeks). (Data source: Worldometer: https://www.worldometers.info/coronavirus/coronavirus/country/india).

A

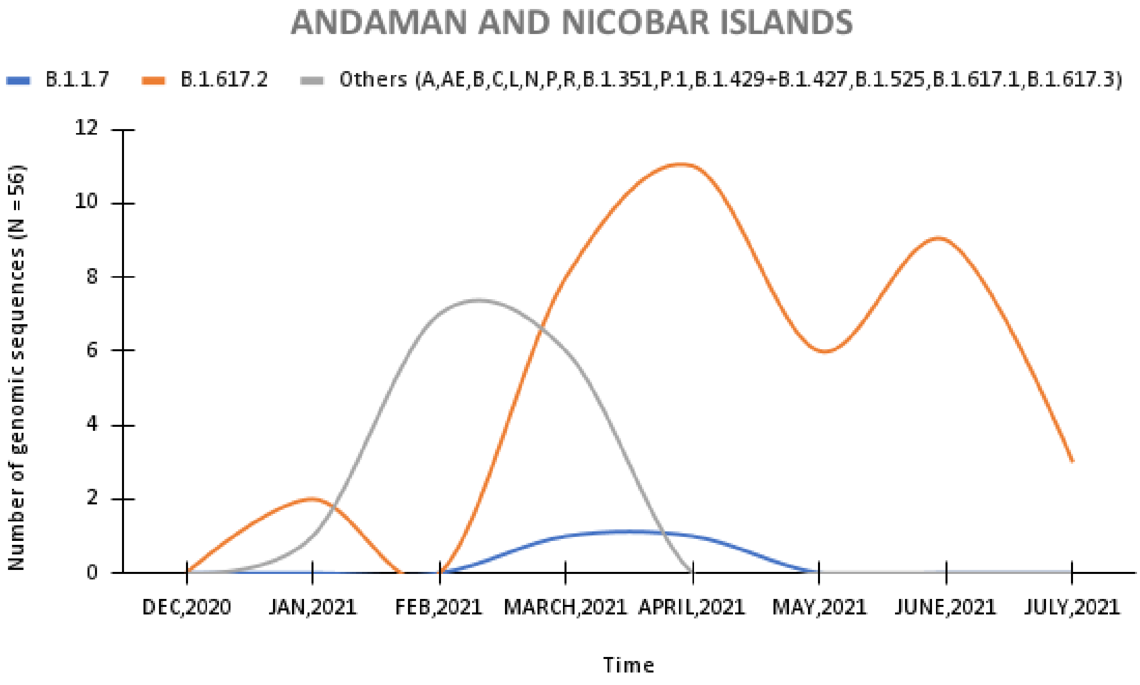
ANDAMAN AND NICOBAR ISLANDS
B.1.1.7
B.1.617.2
Others (A,AE,B,C,L,N,P,R,B.1.351,P.1,B.1.429+B.1.427,B.1.525,B.1.617.1,B.1.617.3)
Number of genomic sequences (N =56)
0
2
4
6
8
10
12
DEC,2020
JAN,2021
FEB,2021
MARCH,2021
APRIL,2021
MAY,2021
JUNE,2021
JULY,2021
Time

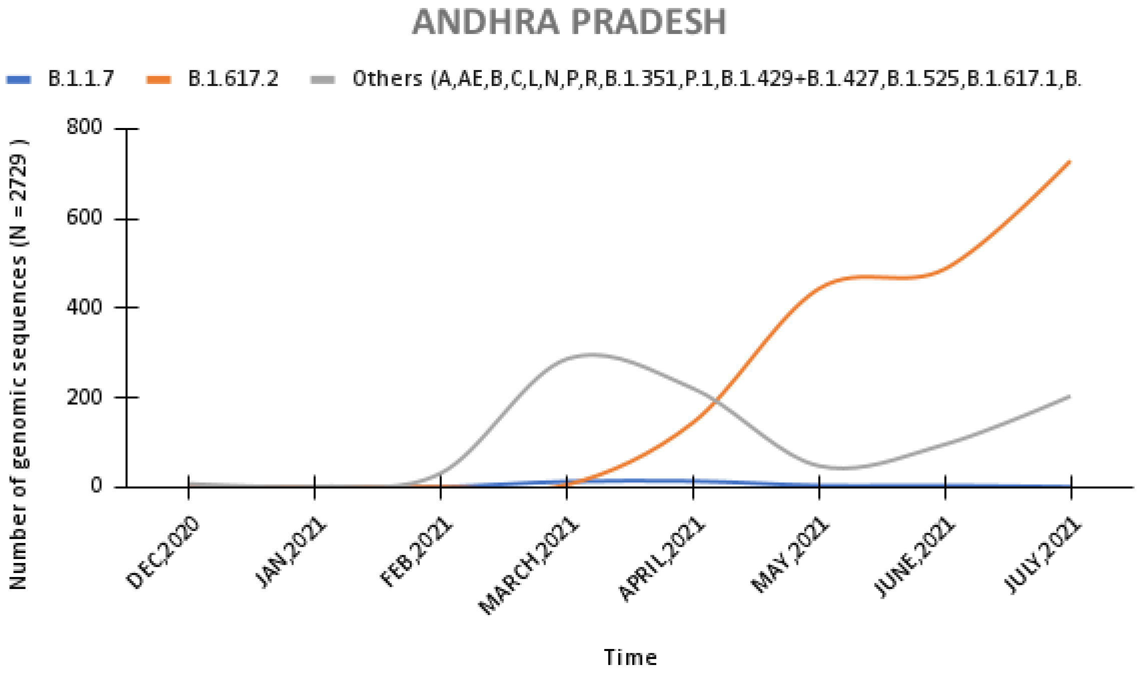
ANDHRA PRADESH
B.1.1.7
B.1.617.2
Others (A,AE,B,C,L,N,P,R,B.1.351,P.1,B.1.429+B.1.427,B.1.525,B.1.617.1,B.
Number of genomic sequences (N = 2729 )
0
200
400
600
800
DEC,2020
JAN,2021
FEB,2021
MARCH,2021
APRIL,2021
MAY,2021
JUNE,2021
JULY,2021
Time

B

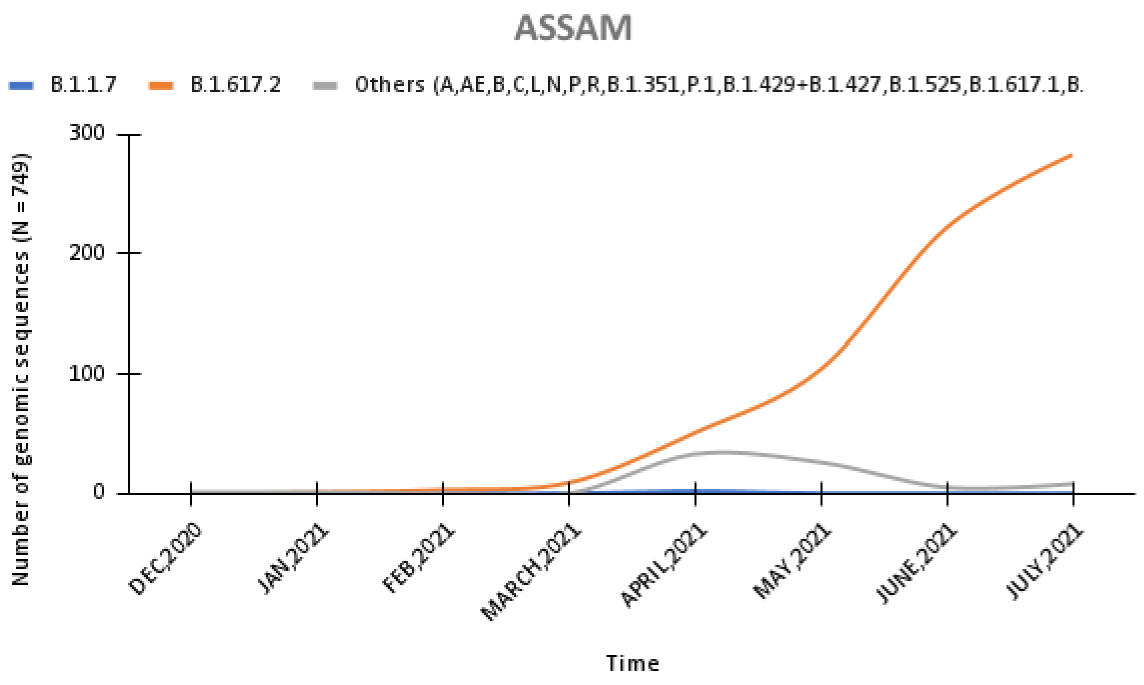
ASSAM
B.1.1.7
B.1.617.2
Others (A,AE,B,C,L,N,P,R,B.1.351,P.1,B.1.429+B.1.427,B.1.525,B.1.617.1,B.
Number of genomic sequences (N = 749)
300
200
100
0
DEC,2020
JAN,2021
FEB,2021
MARCH,2021
APRIL,2021
MAY,2021
JUNE,2021
JULY,2021
Time

C

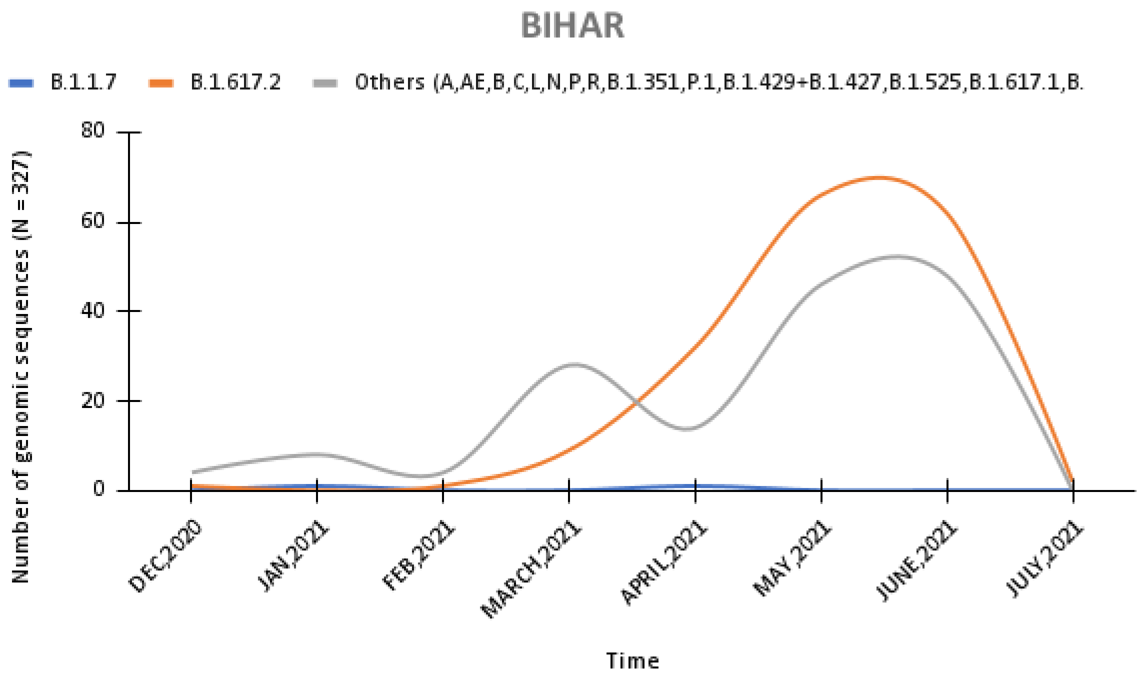
BIHAR
B.1.1.7
B.1.617.2
Others (A,AE,B,C,L,N,P,R,B.1.351,P.1,B.1.429+B.1.427,B.1.525,B.1.617.1,B.
Number of genomic sequences (N = 327)
80
60
40
20
0
DEC,2020
JAN,2021
FEB,2021
MARCH,2021
APRIL,2021
MAY,2021
JUNE,2021
JULY,2021
Time

D

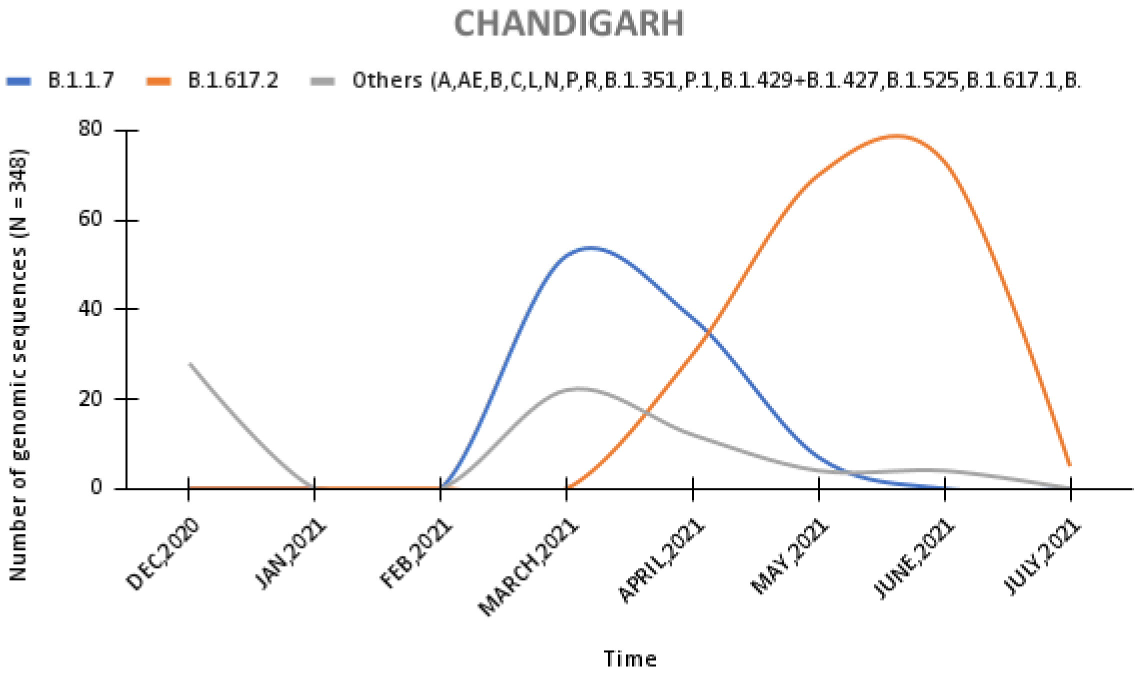
CHANDIGARH
B.1.1.7
B.1.617.2
Others (A,AE,B,C,L,N,P,R,B.1.351,P.1,B.1.429+B.1.427,B.1.525,B.1.617.1,B.
Number of genomic sequences (N = 348)
80
60
40
20
0
DEC,2020
JAN,2021
FEB,2021
MARCH,2021
APRIL,2021
MAY,2021
JUNE,2021
JULY,2021
Time

E

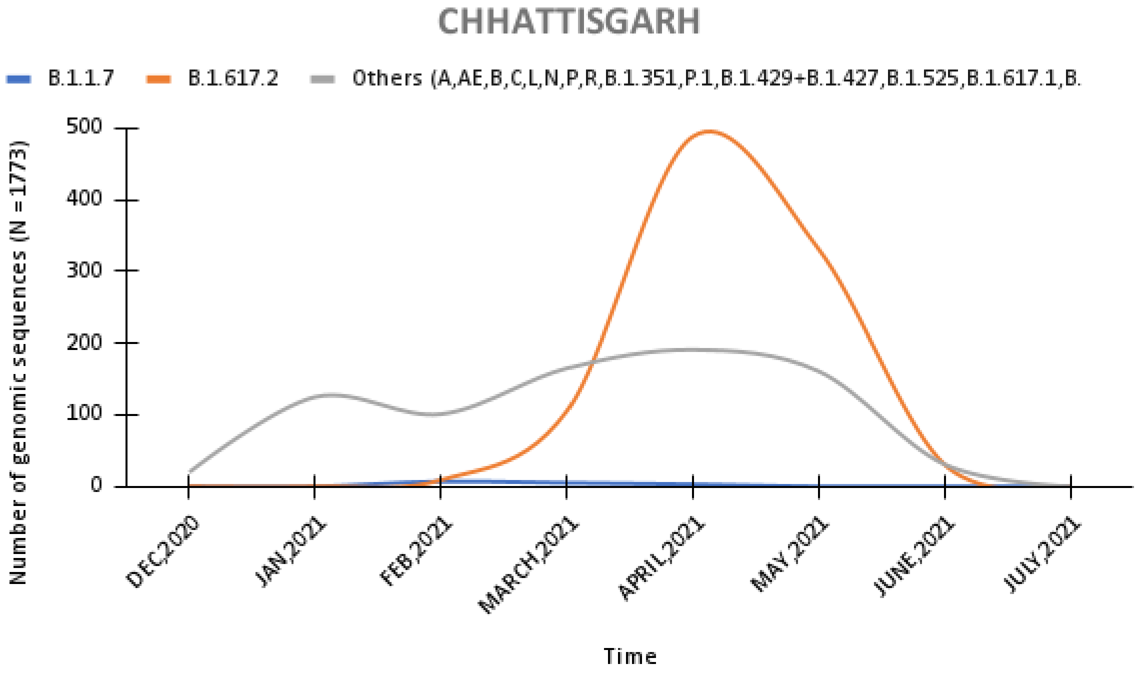
CHHATTISGARH
B.1.1.7
B.1.617.2
Others (A,AE,B,C,L,N,P,R,B.1.351,P.1,B.1.429+B.1.427,B.1.525,B.1.617.1,B.
Number of genomic sequences (N = 1773)
500
400
300
200
100
0
DEC,2020
JAN,2021
FEB,2021
MARCH,2021
APRIL,2021
MAY,2021
JUNE,2021
JULY,2021
Time

F

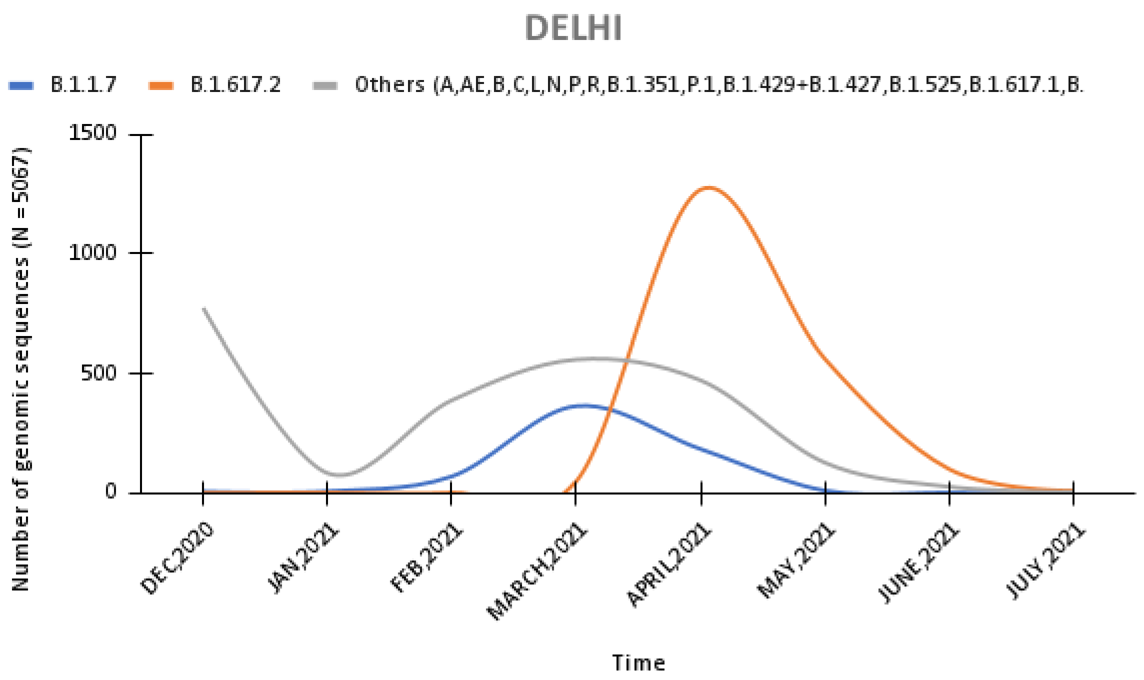
DELHI
B.1.1.7
B.1.617.2
Others (A,AE,B,C,L,N,P,R,B.1.351,P.1,B.1.429+B.1.427,B.1.525,B.1.617.1,B.
Number of genomic sequences (N = 5067)
1500
1000
500
0
DEC,2020
JAN,2021
FEB,2021
MARCH,2021
APRIL,2021
MAY,2021
JUNE,2021
JULY,2021
Time

G

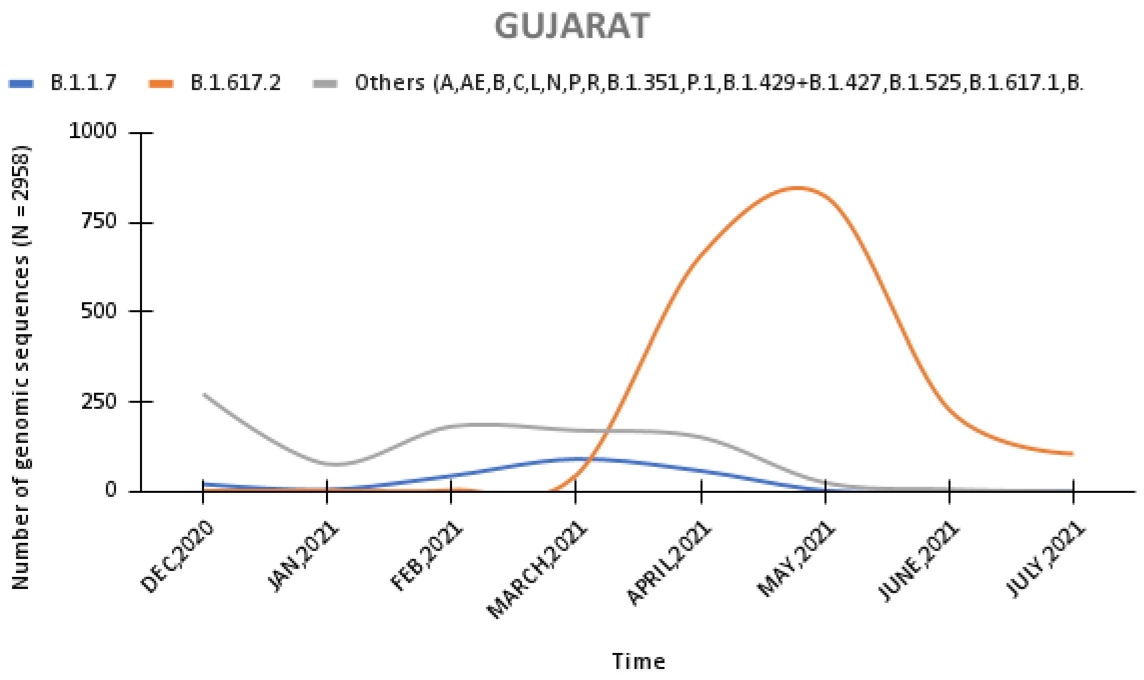
GUJARAT
B.1.1.7
B.1.617.2
Others (A,AE,B,C,L,N,P,R,B.1.351,P.1,B.1.429+B.1.427,B.1.525,B.1.617.1,B.
Number of genomic sequences (N = 2958)
1000
750
500
250
0
DEC,2020
JAN,2021
FEB,2021
MARCH,2021
APRIL,2021
MAY,2021
JUNE,2021
JULY,2021
Time

H

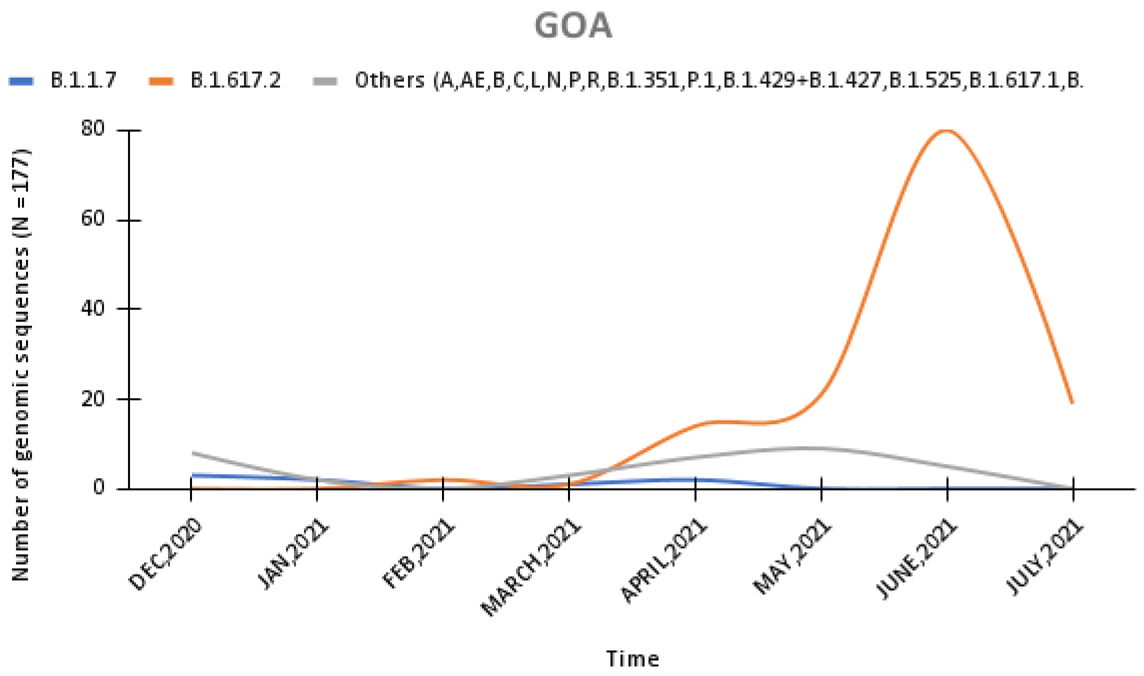
GOA
B.1.1.7
B.1.617.2
Others (A,AE,B,C,L,N,P,R,B.1.351,P.1,B.1.429+B.1.427,B.1.525,B.1.617.1,B.
Number of genomic sequences (N =177)
80
60
40
20
0
DEC,2020
JAN,2021
FEB,2021
MARCH,2021
APRIL,2021
MAY,2021
JUNE,2021
JULY,2021
Time

I

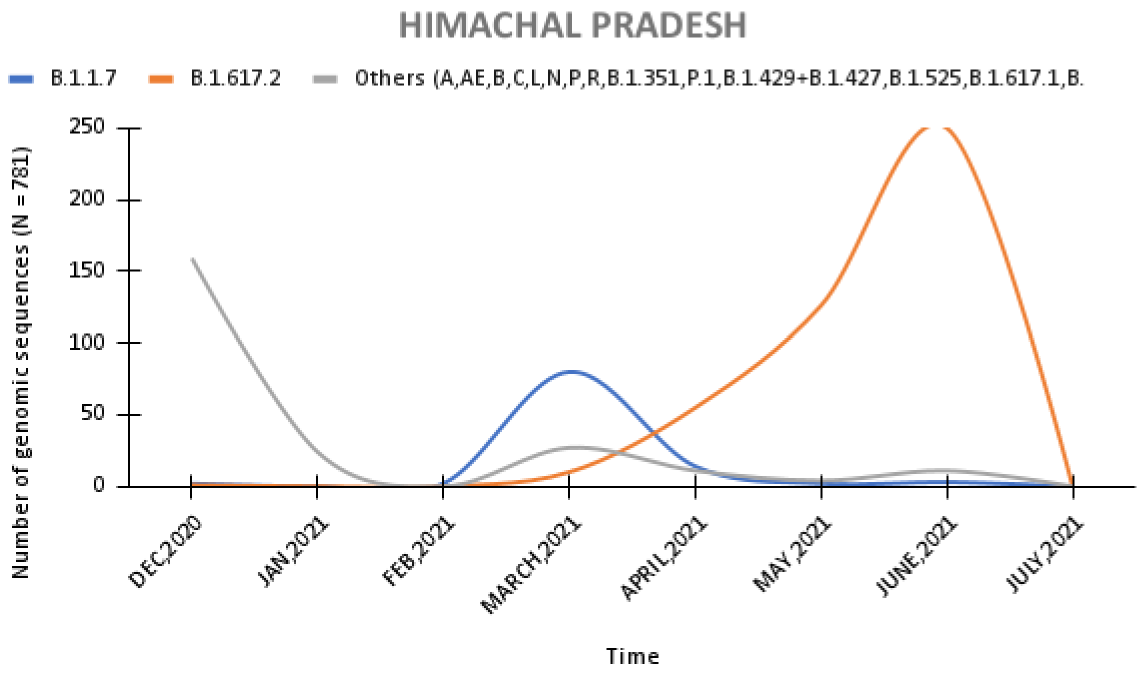
HIMACHAL PRADESH
B.1.1.7
B.1.617.2
Others (A,AE,B,C,L,N,P,R,B.1.351,P.1,B.1.429+B.1.427,B.1.525,B.1.617.1,B.
Number of genomic sequences (N = 781)
250
200
150
100
50
0
DEC,2020
JAN,2021
FEB,2021
MARCH,2021
APRIL,2021
MAY,2021
JUNE,2021
JULY,2021
Time

J

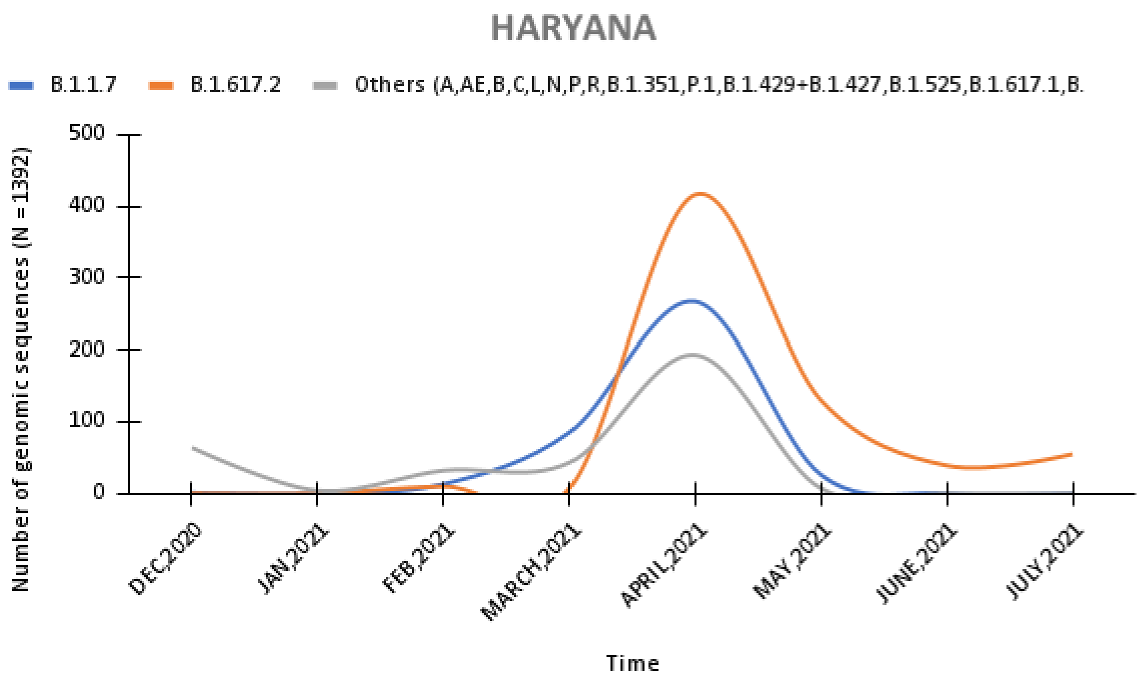
HARYANA
B.1.1.7
B.1.617.2
Others (A,AE,B,C,L,N,P,R,B.1.351,P.1,B.1.429+B.1.427,B.1.525,B.1.617.1,B.
Number of genomic sequences (N =1392)
500
400
300
200
100
0
DEC,2020
JAN,2021
FEB,2021
MARCH,2021
APRIL,2021
MAY,2021
JUNE,2021
JULY,2021
Time

K

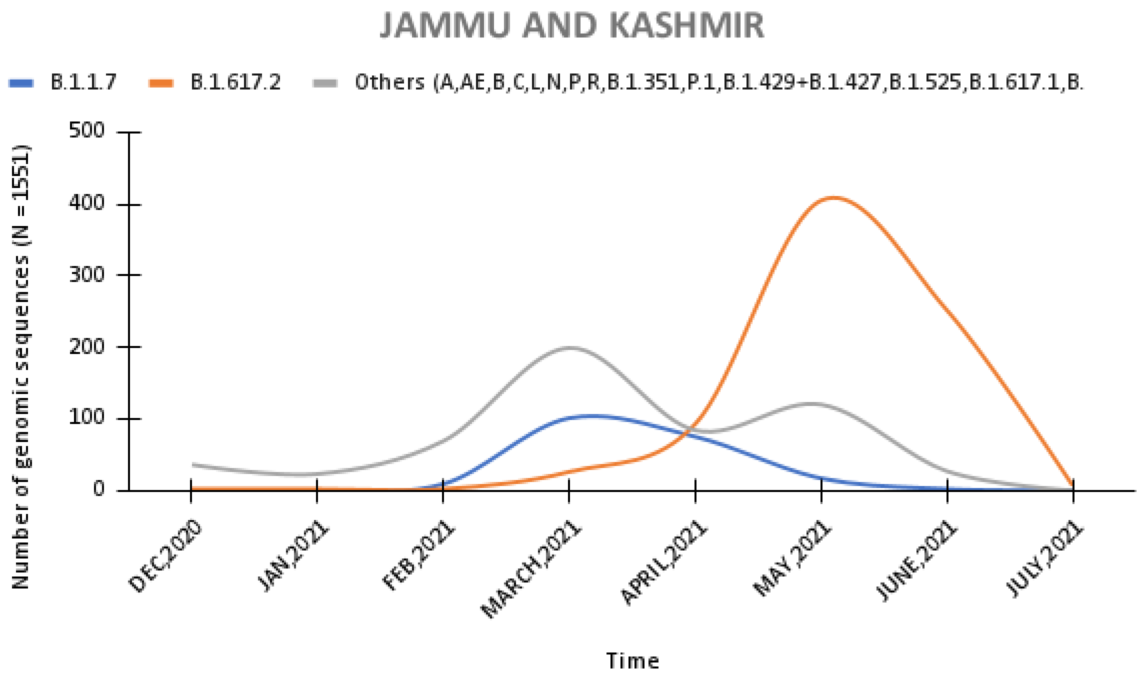
JAMMU AND KASHMIR
B.1.1.7
B.1.617.2
Others (A,AE,B,C,L,N,P,R,B.1.351,P.1,B.1.429+B.1.427,B.1.525,B.1.617.1,B.
Number of genomic sequences (N =1551)
500
400
300
200
100
0
DEC,2020
JAN,2021
FEB,2021
MARCH,2021
APRIL,2021
MAY,2021
JUNE,2021
JULY,2021
Time

L

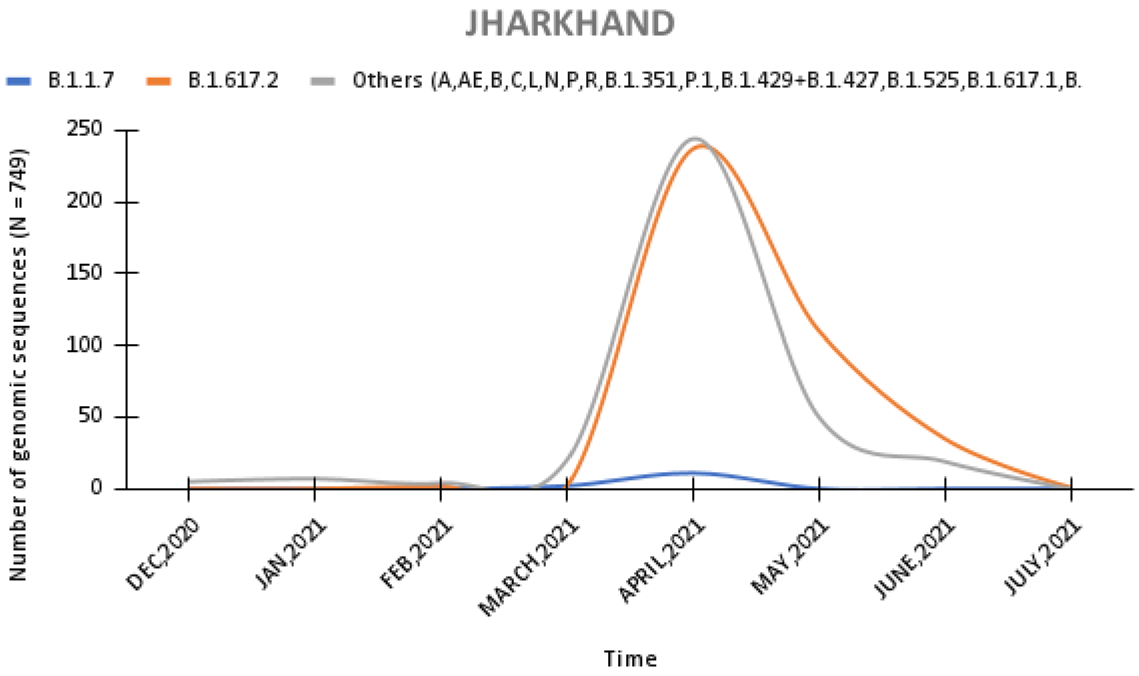
JHARKHAND
B.1.1.7
B.1.617.2
Others (A,AE,B,C,L,N,P,R,B.1.351,P.1,B.1.429+B.1.427,B.1.525,B.1.617.1,B.
Number of genomic sequences (N = 749)
250
200
150
100
50
0
DEC,2020
JAN,2021
FEB,2021
MARCH,2021
APRIL,2021
MAY,2021
JUNE,2021
JULY,2021
Time

M

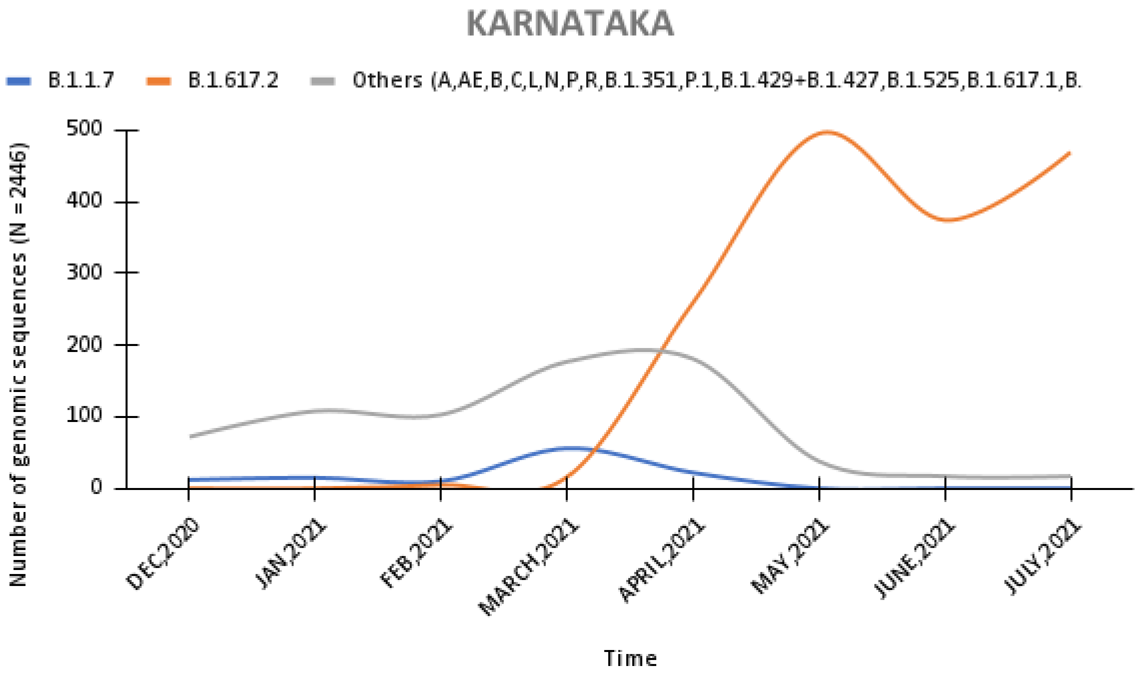
KARNATAKA
B.1.1.7
B.1.617.2
Others (A,AE,B,C,L,N,P,R,B.1.351,P.1,B.1.429+B.1.427,B.1.525,B.1.617.1,B.
Number of genomic sequences (N = 2446)
500
400
300
200
100
0
DEC,2020
JAN,2021
FEB,2021
MARCH,2021
APRIL,2021
MAY,2021
JUNE,2021
JULY,2021
Time

N

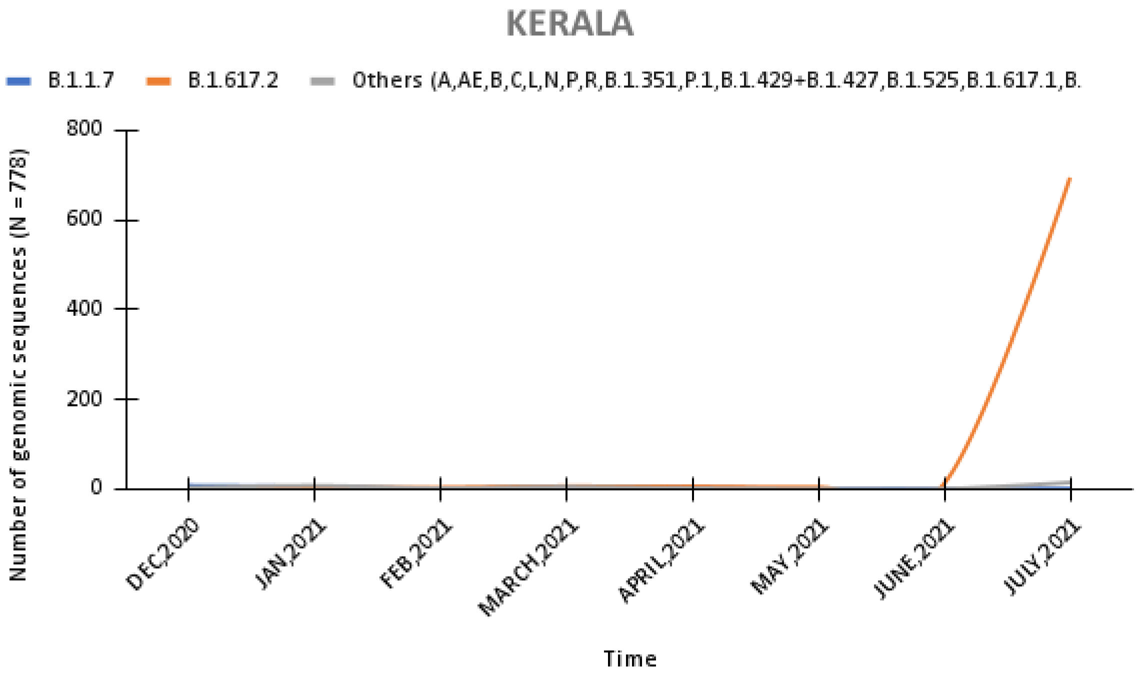
KERALA
B.1.1.7
B.1.617.2
Others (A,AE,B,C,L,N,P,R,B.1.351,P.1,B.1.429+B.1.427,B.1.525,B.1.617.1,B.
Number of genomic sequences (N = 778)
800
600
400
200
0
DEC,2020
JAN,2021
FEB,2021
MARCH,2021
APRIL,2021
MAY,2021
JUNE,2021
JULY,2021
Time

O

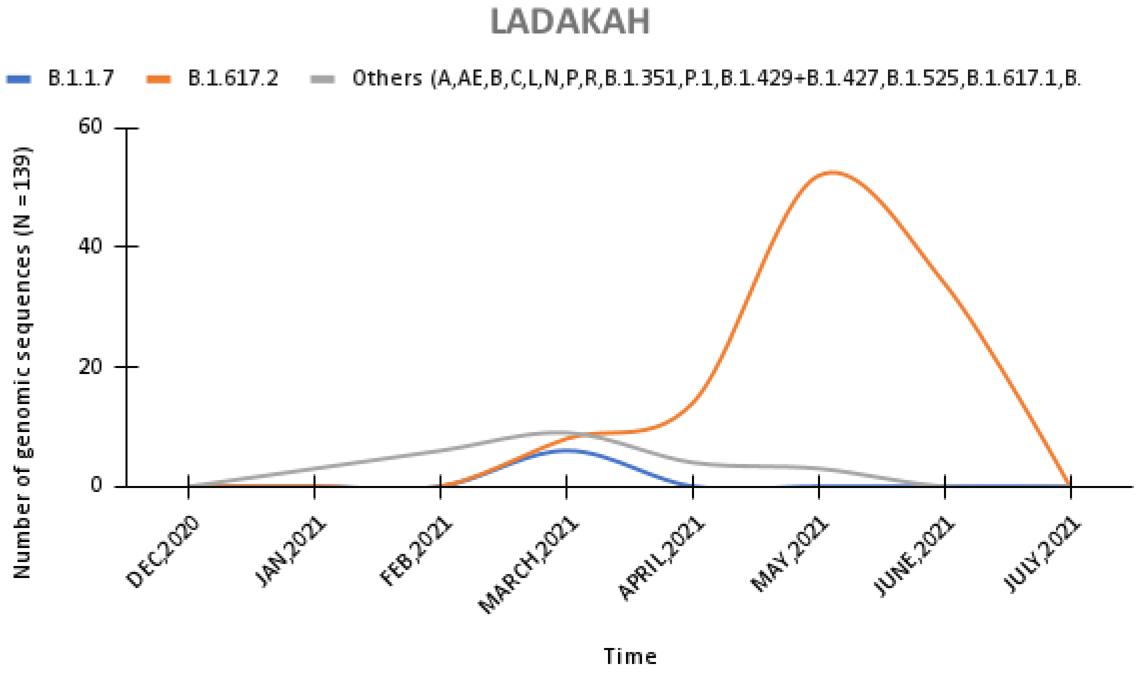
LADAKAH
B.1.1.7
B.1.617.2
Others (A,AE,B,C,L,N,P,R,B.1.351,P.1,B.1.429+B.1.427,B.1.525,B.1.617.1,B.
Number of genomic sequences (N = 139)
60
40
20
0
DEC,2020
JAN,2021
FEB,2021
MARCH,2021
APRIL,2021
MAY,2021
JUNE,2021
JULY,2021
Time

P

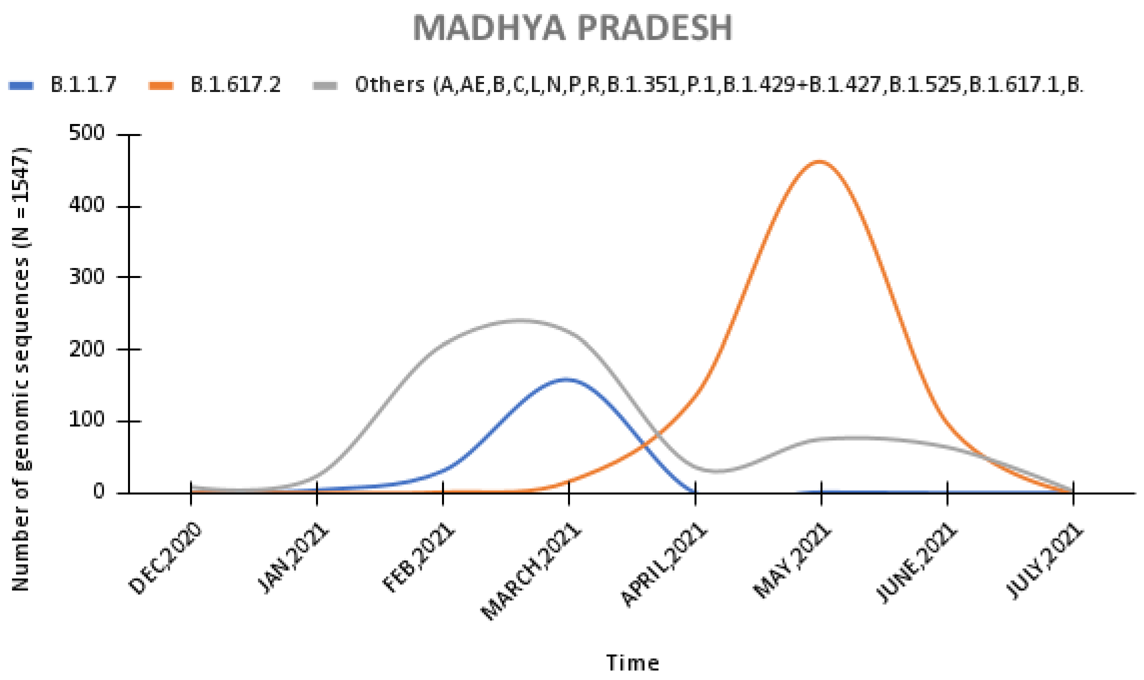
MADHYA PRADESH
B.1.1.7
B.1.617.2
Others (A,AE,B,C,L,N,P,R,B.1.351,P.1,B.1.429+B.1.427,B.1.525,B.1.617.1,B.
Number of genomic sequences (N = 1547)
500
400
300
200
100
0
DEC,2020
JAN,2021
FEB,2021
MARCH,2021
APRIL,2021
MAY,2021
JUNE,2021
JULY,2021
Time

Q

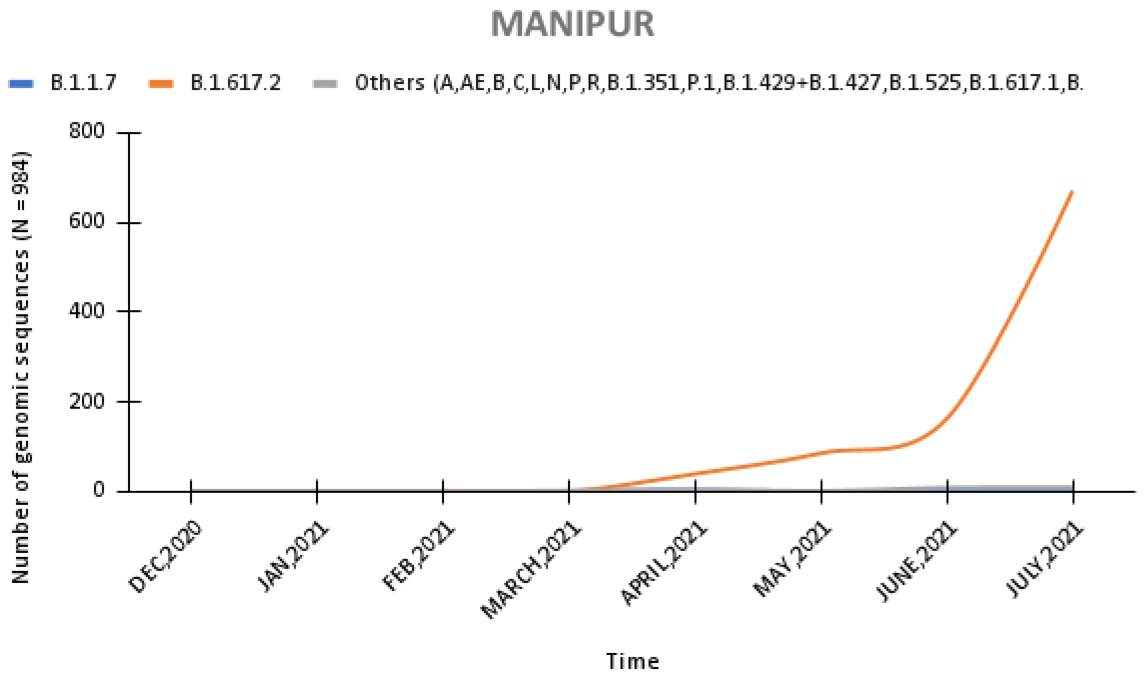
MANIPUR
B.1.1.7
B.1.617.2
Others (A,AE,B,C,L,N,P,R,B.1.351,P.1,B.1.429+B.1.427,B.1.525,B.1.617.1,B.
Number of genomic sequences (N = 984)
800
600
400
200
0
DEC,2020
JAN,2021
FEB,2021
MARCH,2021
APRIL,2021
MAY,2021
JUNE,2021
JULY,2021
Time

R

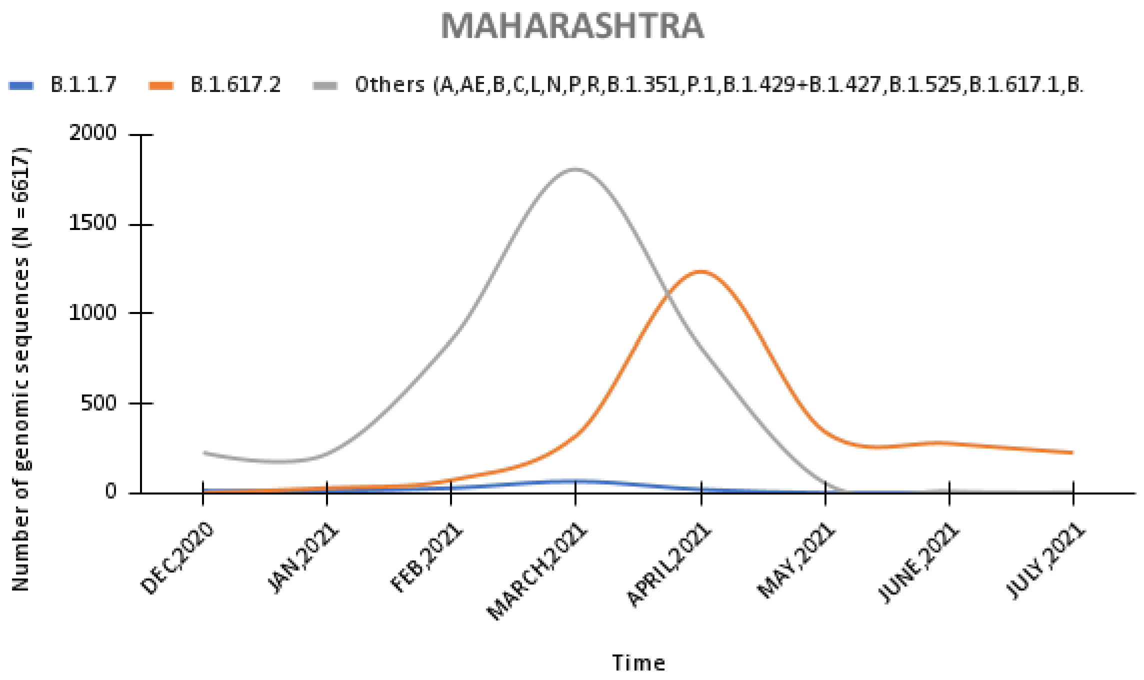
MAHARASHTRA
B.1.1.7
B.1.617.2
Others (A,AE,B,C,L,N,P,R,B.1.351,P.1,B.1.429+B.1.427,B.1.525,B.1.617.1,B.
Number of genomic sequences (N = 6617)
2000
1500
1000
500
0
DEC,2020
JAN,2021
FEB,2021
MARCH,2021
APRIL,2021
MAY,2021
JUNE,2021
JULY,2021
Time

S

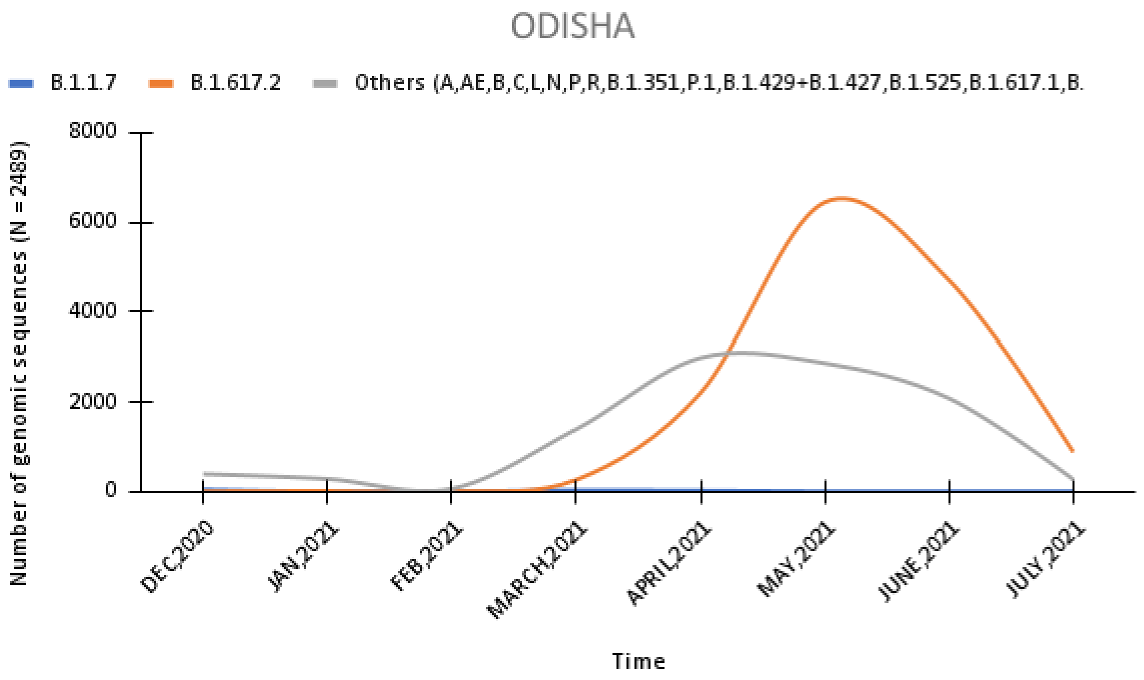
ODISHA
B.1.1.7
B.1.617.2
Others (A,AE,B,C,L,N,P,R,B.1.351,P.1,B.1.429+B.1.427,B.1.525,B.1.617.1,B.
Number of genomic sequences (N = 2489)
8000
6000
4000
2000
0
DEC,2020
JAN,2021
FEB,2021
MARCH,2021
APRIL,2021
MAY,2021
JUNE,2021
JULY,2021
Time

T

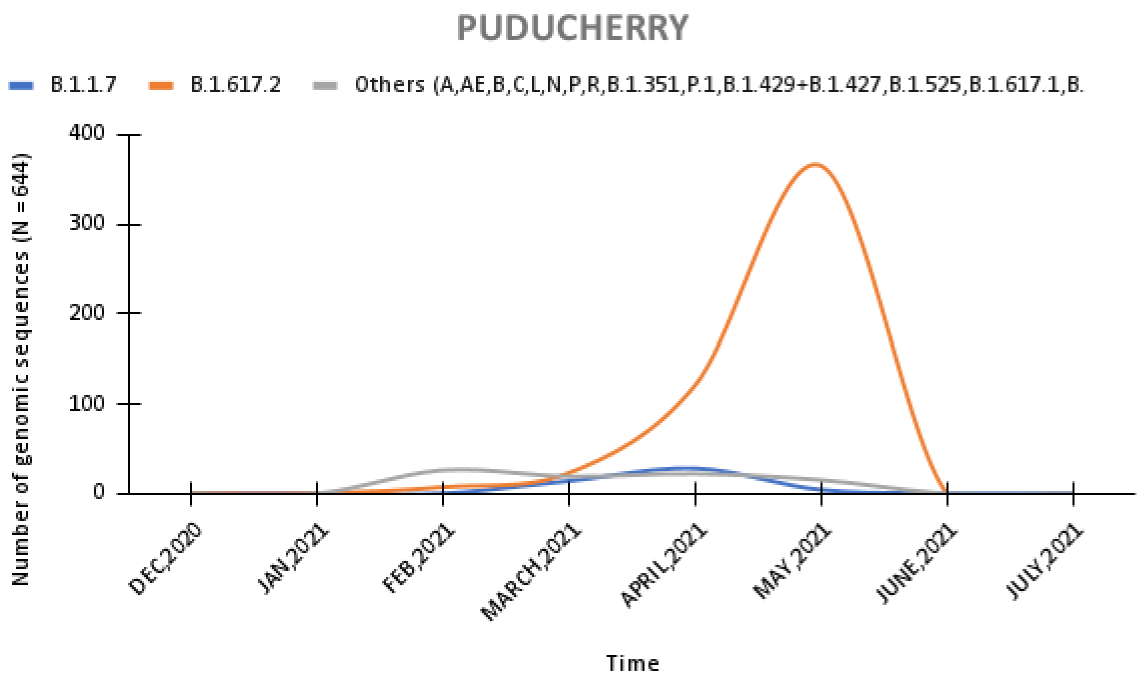

PUDUCHERRY
B.1.1.7
B.1.617.2
Others (A,AE,B,C,L,N,P,R,B.1.351,P.1,B.1.429+B.1.427,B.1.525,B.1.617.1,B.
Number of genomic sequences (N =644)
400
300
200
100
0
DEC,2020
JAN,2021
FEB,2021
MARCH,2021
APRIL,2021
MAY,2021
JUNE,2021
JULY,2021
Time


U

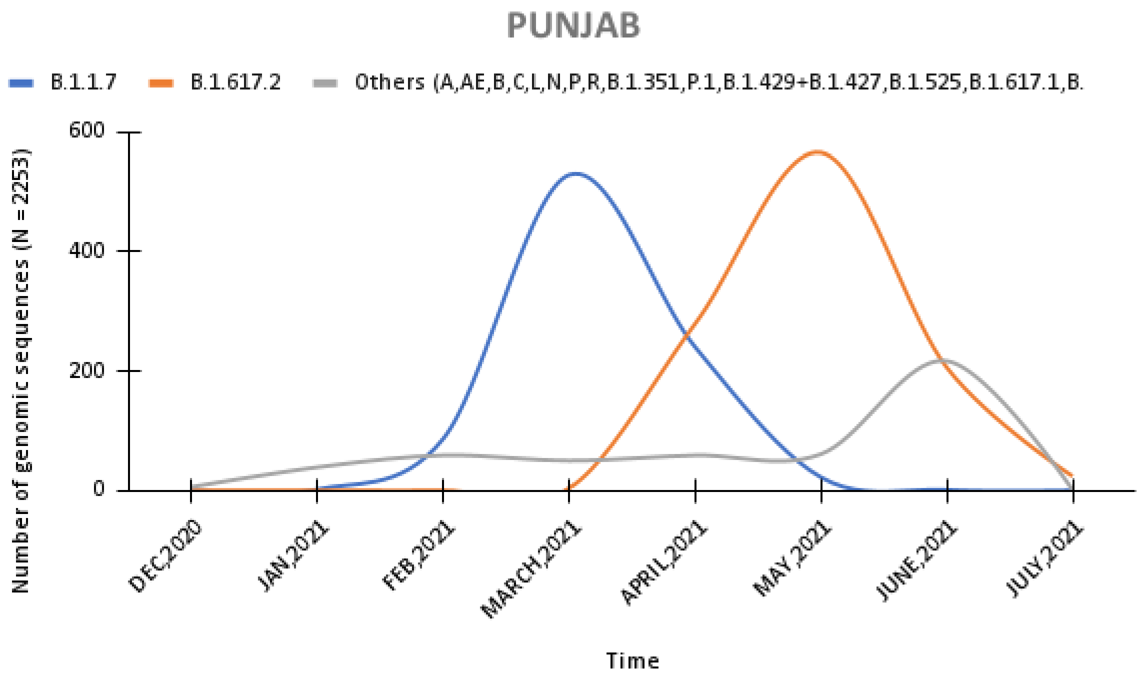

PUNJAB
B.1.1.7
B.1.617.2
Others (A,AE,B,C,L,N,P,R,B.1.351,P.1,B.1.429+B.1.427,B.1.525,B.1.617.1,B.
Number of genomic sequences (N = 2253)
600
400
200
0
DEC,2020
JAN,2021
FEB,2021
MARCH,2021
APRIL,2021
MAY,2021
JUNE,2021
JULY,2021
Time


V

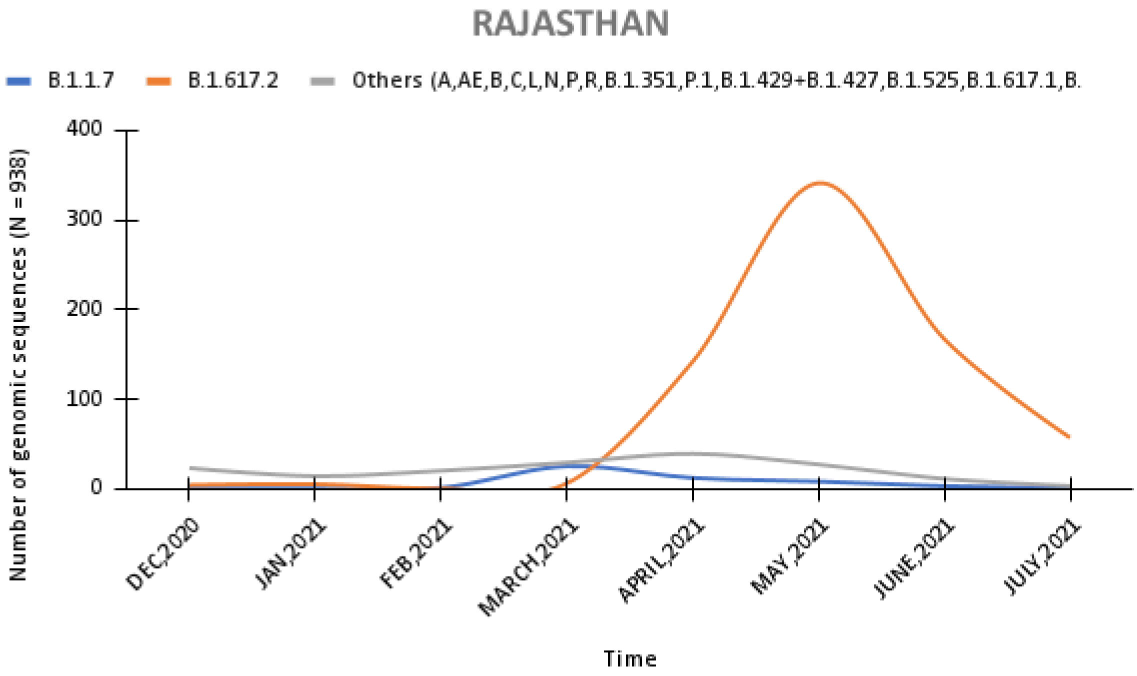
RAJASTHAN
B.1.1.7
B.1.617.2
Others (A,AE,B,C,L,N,P,R,B.1.351,P.1,B.1.429+B.1.427,B.1.525,B.1.617.1,B.
Number of genomic sequences (N =938)
400
300
200
100
0
DEC,2020
JAN,2021
FEB,2021
MARCH,2021
APRIL,2021
MAY,2021
JUNE,2021
JULY,2021
Time

W

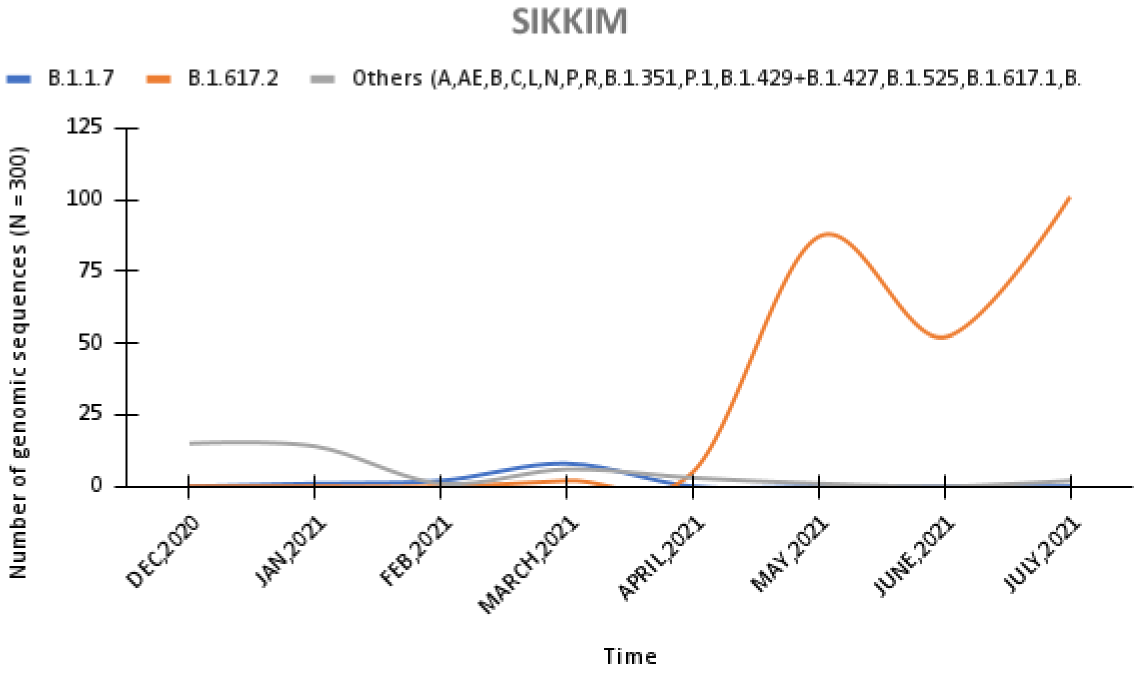
SIKKIM
B.1.1.7
B.1.617.2
Others (A,AE,B,C,L,N,P,R,B.1.351,P.1,B.1.429+B.1.427,B.1.525,B.1.617.1,B.
Number of genomic sequences (N =300)
125
100
75
50
25
0
DEC,2020
JAN,2021
FEB,2021
MARCH,2021
APRIL,2021
MAY,2021
JUNE,2021
JULY,2021
Time

X

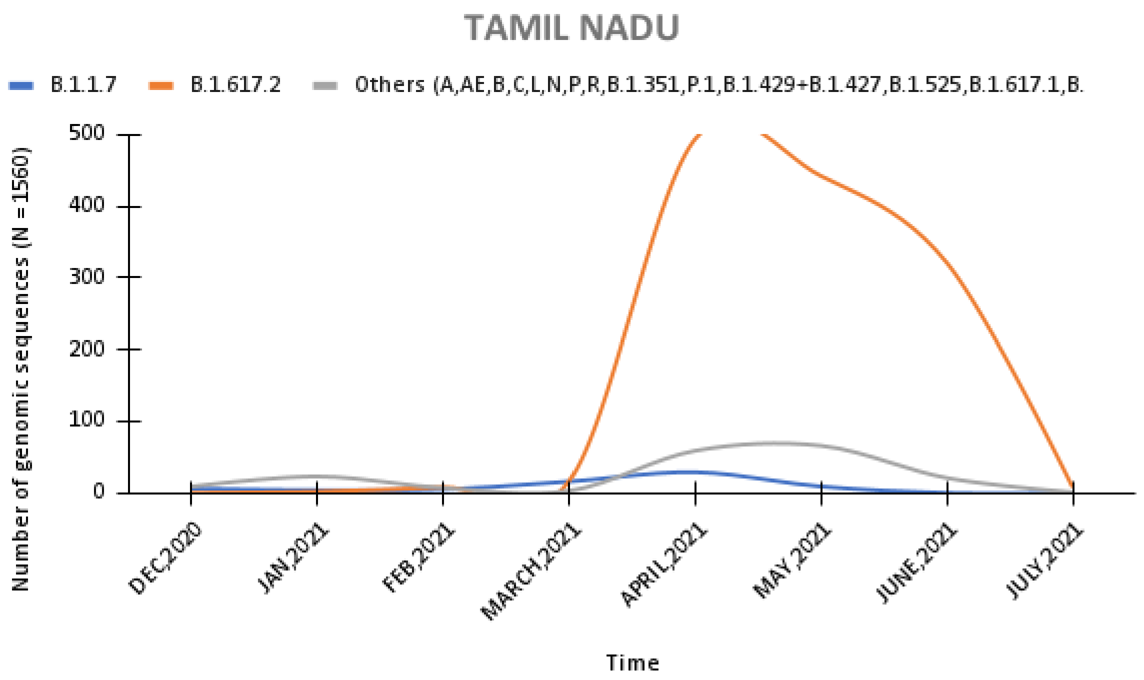
TAMIL NADU
B.1.1.7
B.1.617.2
Others (A,AE,B,C,L,N,P,R,B.1.351,P.1,B.1.429+B.1.427,B.1.525,B.1.617.1,B.
Number of genomic sequences (N = 1560)
500
400
300
200
100
0
DEC,2020
JAN,2021
FEB,2021
MARCH,2021
APRIL,2021
MAY,2021
JUNE,2021
JULY,2021
Time

Y

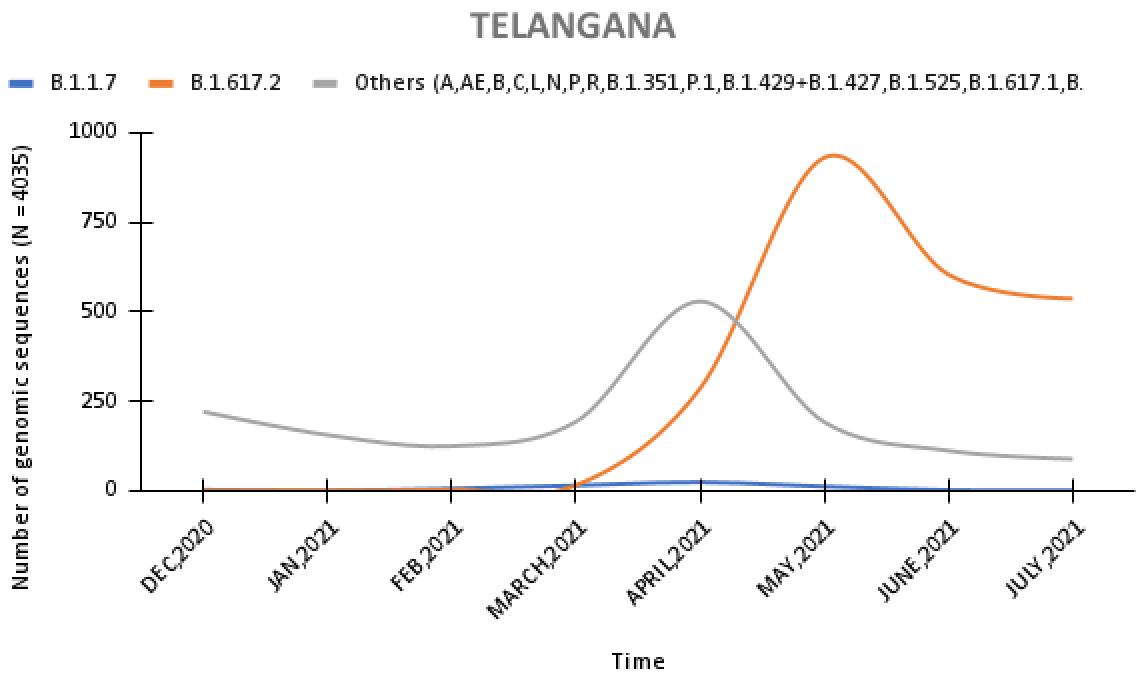
TELANGANA
B.1.1.7
B.1.617.2
Others (A,AE,B,C,L,N,P,R,B.1.351,P.1,B.1.429+B.1.427,B.1.525,B.1.617.1,B.
Number of genomic sequences (N = 4035)
1000
750
500
250
0
DEC,2020
JAN,2021
FEB,2021
MARCH,2021
APRIL,2021
MAY,2021
JUNE,2021
JULY,2021
Time

Z

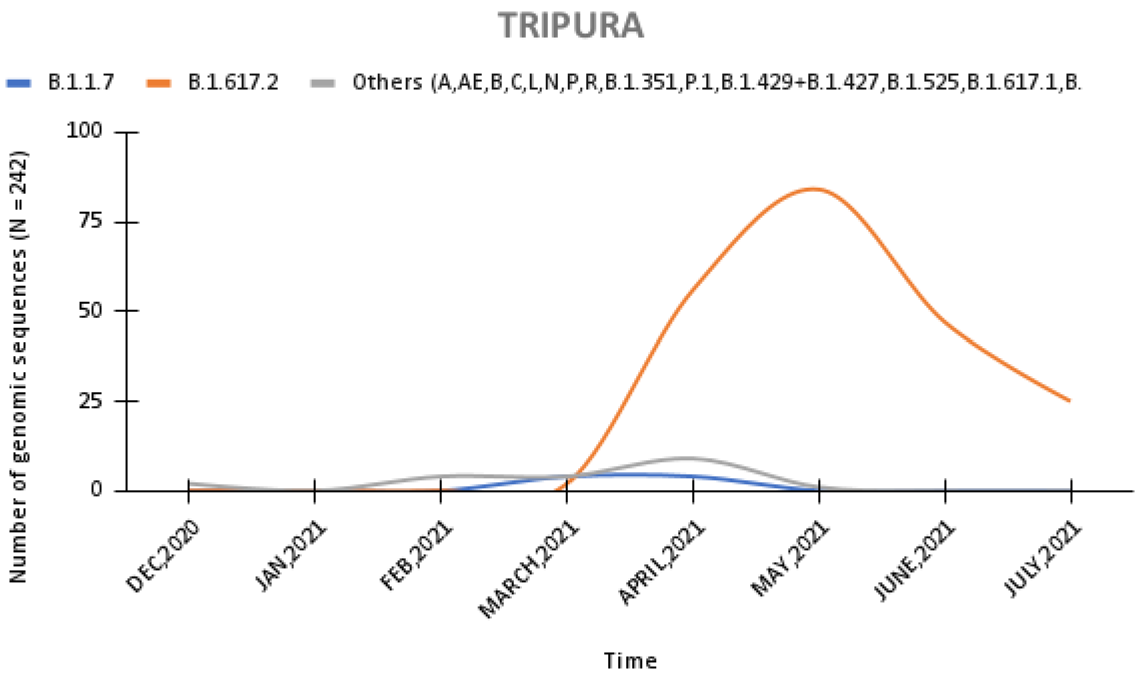
TRIPURA
B.1.1.7
B.1.617.2
Others (A,AE,B,C,L,N,P,R,B.1.351,P.1,B.1.429+B.1.427,B.1.525,B.1.617.1,B.
Number of genomic sequences (N = 242)
100
75
50
25
0
DEC,2020
JAN,2021
FEB,2021
MARCH,2021
APRIL,2021
MAY,2021
JUNE,2021
JULY,2021
Time

AA

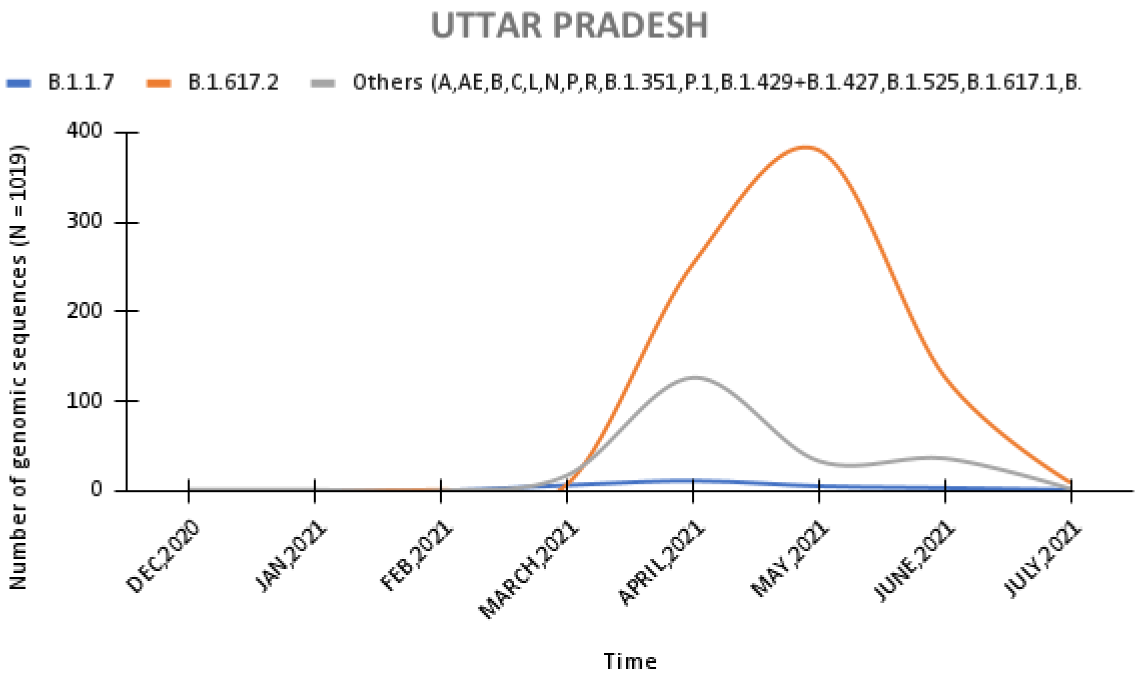
UTTAR PRADESH
B.1.1.7
B.1.617.2
Others (A,AE,B,C,L,N,P,R,B.1.351,P.1,B.1.429+B.1.427,B.1.525,B.1.617.1,B.
Number of genomic sequences (N = 1019)
400
300
200
100
0
DEC,2020
JAN,2021
FEB,2021
MARCH,2021
APRIL,2021
MAY,2021
JUNE,2021
JULY,2021
Time

AB

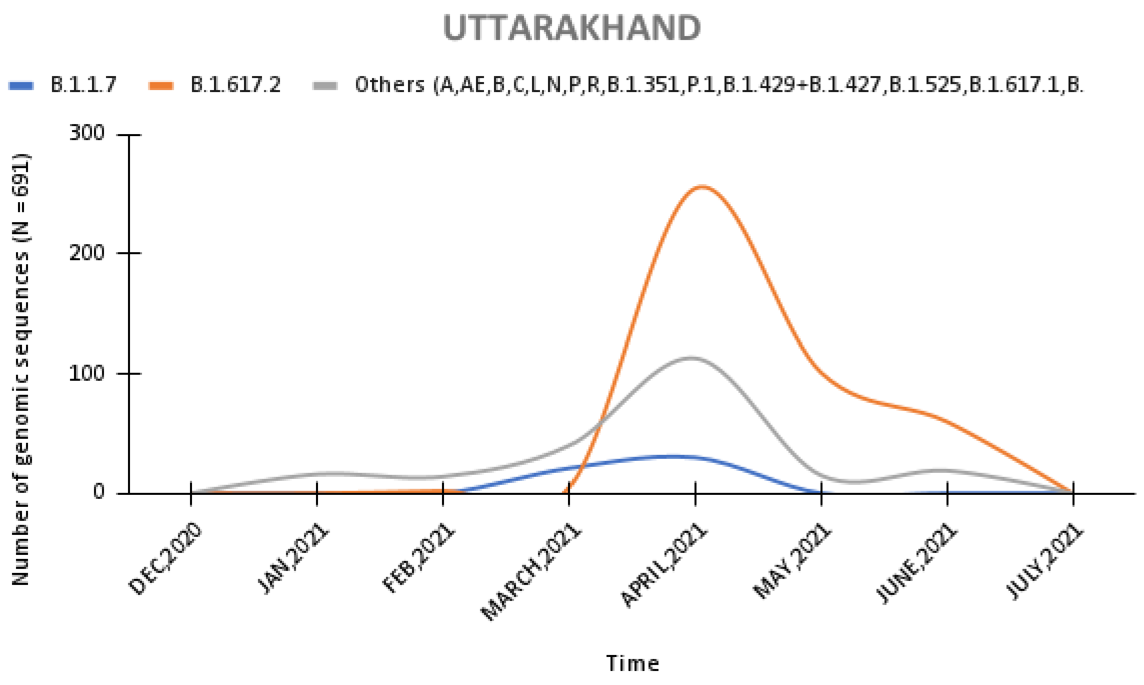


AC

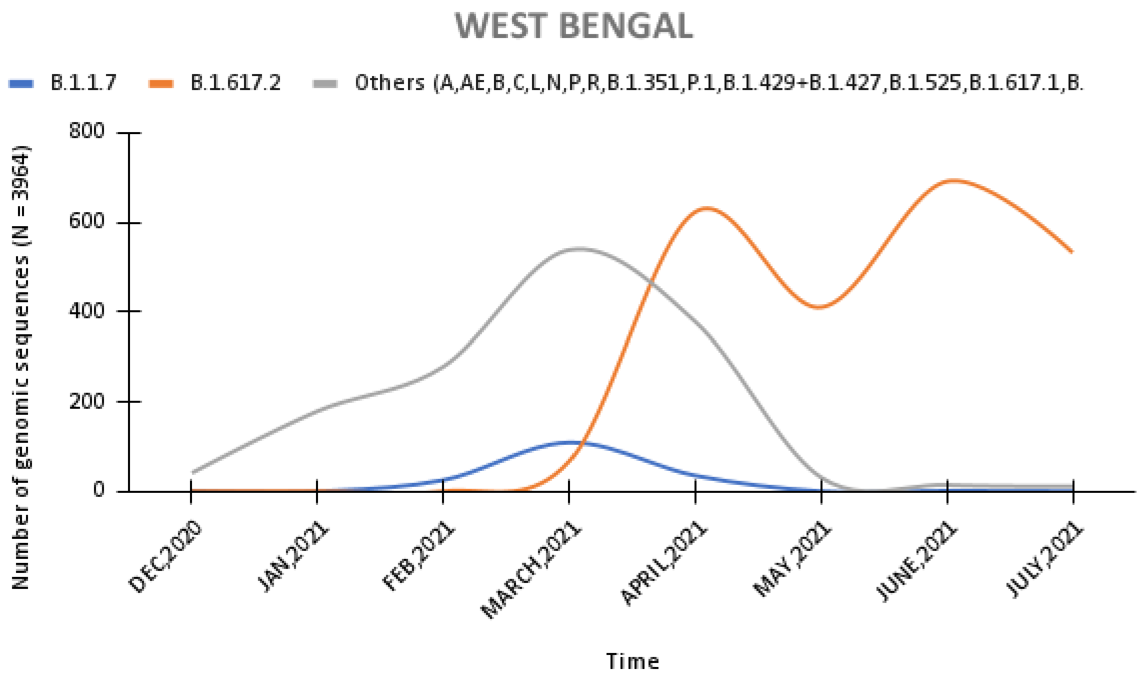


AD

**Figure S2 Monthly distribution of SARS-CoV-2 variants in genomic sequence data from states and union territories of India uploaded on GISAID database for the period of 1st December 2020 to 26th July 2021.** Data source: SARS-CoV-2 genomic sequence—GISAID database: https://www.gisaid.org.
